# Addressing selection bias in the UK Biobank neurological imaging cohort

**DOI:** 10.1101/2022.01.13.22269266

**Authors:** Valerie Bradley, Thomas E. Nichols

**Affiliations:** Department of Statistics, University of Oxford; Big Data Institute, Li Ka Shing Centre for Health Information and Discovery, Nuffield Department of Population Health, University of Oxford; Wellcome Centre for Integrative Neuroimaging, FMRIB, Nuffield Department of Clinical Neurosciences, University of Oxford

## Abstract

The UK Biobank is a national prospective study of half a million participants between the ages of 40 and 69 at the time of recruitment between 2006 and 2010, established to facilitate research on diseases of aging. The imaging cohort is a subset of UK Biobank participants who have agreed to undergo extensive additional imaging assessments. However, Fry et al. (2017) finds evidence of “healthy volunteer bias” in the UK Biobank – participants are less likely to smoke, be obese, consume alcohol daily than the target population of UK adults. Here we examine selection bias in the UK Biobank imaging cohort. We address two common misconceptions: first, that study size can compensate for bias in data collection, and second that selection bias does not affect estimates of associations, which are the primary interest of the UK Biobank. We introduce inverse probability weighting (IPW) as an approach commonly used in survey research that can be used to address selection bias in volunteer health studies like the UK Biobank. We discuss 6 such methods – five existing and one novel –, assess relative performance in simulation studies, and apply them to the UK Biobank imaging cohort. We find that our novel method, BART for predicting the probability of selection combined with raking, performs well relative to existing methods, and helps alleviate selection bias in the UK Biobank imaging cohort.

## 1 Introduction

The UK Biobank (UKB) is a national prospective study of half a million participants between the ages of 40 and 69 at the time of recruitment between 2006 and 2010. The UK Biobank was established to examine relationships between exposures and common health-related outcomes that affect aging populations, for example cancer, heart disease, diabetes, and dementia (Sudlow et al., 2015). Though the study design took steps to maximize the generalizability of the UKB cohort, recruiting enough participants for analysis of complex exposure-outcome relationships was of greater concern (Sudlow et al., 2015). As a result, Fry et al. (2017) describes how the cohort suffers from “healthy volunteer” bias, in that participants exhibit lower rates of smoking, obesity, and daily alcohol consumption than the target population of UK adults. Strikingly, Fry et al. (2017) note that “all-cause mortality is approximately half that of the UK population as a whole, and total cancer incidence rates are approximately 10%-20% lower.”

This healthy volunteer bias is a form of selection bias – when observed data is not representative of the population of interest due to, for example, self-selection of participants or analysis decisions. (J.Heckman, 1979; Little and Rubin, 1986). Selection bias threatens the generalizability of study estimates to other populations. Despite the evidence of selection bias in the UKB, Fry et al. (2017) conclude that “Although UK Biobank is not suitable for deriving generalizable disease prevalence and incidence rates, its large size and heterogeneity of exposure measures provide valid scientific inferences of associations between exposures and health conditions that are generalizable to other populations.”

However, Meng (2018) proves mathematically that this conclusion is flawed. Large sample sizes cannot (efficiently) overcome bias in data collection, and in fact that a larger sample size can increase overconfidence in estimates when there is bias in data collection – the Big Data Paradox. Bradley et al. (2021) further demonstrates how this Big Data Paradox can even affect large surveys that do take steps to counteract selection bias.

Meng (2018) and Bradley et al. (2021) focus on bias in point estimates of population means, however selection bias (in the form of “collider bias”) is well-known to also affect estimates of associations (Munafó et al., 2018), calling into question the conclusion from Fry et al. (2017) that estimates of associations using the UKB are still valid in the presence of selection bias. LeWinn et al. (2017) demonstrates the impact of selection bias on a sample of 1,162 structural brain images from a community-based sample of children, and find that adjusting for observed selection bias changes estimates of the relationship between age and brain structure.

The UK Biobank is in the process of recruiting a subset of the total 500,000 UK Biobank participants to undergo additional assessments as part of the world’s largest ever multi-modal imaging study (Littlejohns et al., 2020). From the time recruitment began in 2016 to March 2020, over 50,000 participants completed the additional imaging screenings, with the goal of completing 100,000 in total by 2023. Due to the additional respondent burden required to undergo the extensive imaging necessary to participate in this cohort, there is large potential for the selection bias in the UKB to be exacerbated in the imaging cohort.

This paper seeks to quantify the selection bias in the UKB imaging cohort, and to present a set of methods commonly used in survey analysis that could be applied to the UKB to lessen the impact of selection bias on estimates. Section 2 further outlines the UKB imaging cohort data, and introduces the Health Survey for England, which is used to define the target population. Section 3 presents six methods (a mix of standard and novel) that may be used to adjust for selection bias, outlines simulation studies designed to test the relative performance of the proposed methods, and describes how we evaluate the methods’ performances. Results from the selection bias analysis, simulation studies, and application of methods to the UKB imaging cohort are given in Section 4.

## 2 Data

The UK Biobank is a national prospective health study of UK adults ages 40-69 at time of recruitment, which occurred between 2006 and 2010. All participants completed extensive questionnaires, underwent physical and mental health examinations, gave biological samples and consented to have their National Health Service (NHS) records accessed by the study. The goal of the study is to collect data on diseases of aging, before onset Fry et al. (2017).

Additionally, up to 100,000 participants will undergo imaging assessments, including MRI of the brain, heart and abdomen, and full-body bone and joint X-ray. The first subjects were imaged in 2016, and imaging is expected to continue through 2022 (Miller et al., 2016). To date, 34,890 subjects have undergone brain MRI imaging. We restrict our sample to the 20,827 observations that contain complete measurements of T1 total brain volume (grey and white matter), normalized for head size.

To assess selection bias in the UKB imaging cohort, we compare demographic distributions participants to those from the 2016 Health Survey for England (HSE), a high-quality, national health survey. The Health Survey for England (HSE) is an annual survey conducted by the Joint Health Surveys Unit of NatCen Social Research and the Department of Epidemiology and Public Health at University College London (HSE, 2018). We use the 2016 data as it was the latest available at the time this analysis was conducted. The UK Biobank imaging study began in 2016, so there is a slight mismatch in time of collection between the UK Biobank data and our target population, and weighted estimates will correspond to a nationally representative 2016 adult population.

The 2016 HSE interviewed 8,011 adults aged 16 and over, and 2,056 children under the age of 16. We restrict our data to the 4,318 adults aged 44-79 as the UK Biobank imaging subjects only fall within that age range. The health metrics that we are interested in comparing to the UK Biobank are only available for a subset of the overall sample, so we further restrict the sample to 2,348 individuals who are aged 44-79 and underwent a nurse interview. We use the HSE-supplied survey weights for the nurse interview subset for all population calculations. The UK Data Service releases anonymized individual-level results for the HSE, which we use here.

Additional details about the two data sources, as well as an overview of coding and analysis decisions can be found in Appendix A.

## 3 Methods

This section outlines the four sets of methods used. First we discuss the six proposed adjustment methods, then outline how the simulations studies were designed, next discuss how we evaluated the performance of the various methods, and finally give a brief summary of how the methods were applied to the UKB imaging cohort. More detailed descriptions of each method can be found in the Appendix.

### 3.1 Adjustment Methods

We use the structural causal model (SCM) notation to describe the general task of adjusting an estimate for an outcome *Y* in the presence of selection bias (Pearl, 1995b,a; Bareinboim et al., 2014; Bareinboim and Pearl, 2016). This is in contrast to the alternative potential outcomes framework for evaluating missing data (Rubin, 1976; Little and Rubin, 1986).

It is possible to recover an unbiased estimate of the association between **Y** and **X** in the presence of selection bias, if the conditional probability *P* (**y**|**x**) can be expressed as follows in terms of observed quantities:

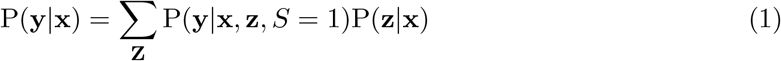

*S* is the selection indicator, equal to 1 if a unit was observed in the sample, and 0 otherwise. **Z** is an *admissible set* of auxiliary variables that blocks paths in the causal graph between **Y** and *S*, such that **Y** ⊥ *S*|**X, Z**. It is possible to recover an unbiased estimate of *P* (**y**|**x** if and only if there is a set **Z** that induces conditional independence between **Y** and *S*, and all the elements of **Z** are observed in the sample and the population. In other words, P(**y**|**x, z**, *S* = 1) and P(**z**|**x**) must be observable.

It is often the case that we don’t observe *P* (**z**|**x**) in the population, because we only observe **Z** in the sample. In this case, to recover *P* (**y**|**x**), we must assume that **Y ⋃ X** ⊥ *S*|**Z**. In this case, we can express the association as

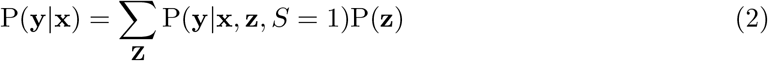

All the methods for adjusting for selection bias that we consider here are Inverse Probability Weight (IPW) techniques (Horvitz and Thompson, 1952). IPW techniques assign weights to each observation in the sample such that – in theory – the weights adjust for unequal probability of selection, such that the weighted sample is representative of the target population. Hernan et al. (2004) describes IPW, when applied correctly, as “creating a pseudopopulation” that produces estimates that are “unaffected by selection bias.”

The motivation for this approach can be seen exactly by re-expressing Equation 2 (Correa et al., 2018):

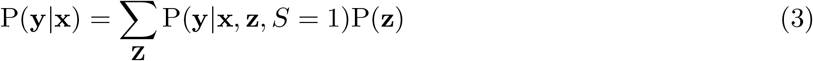

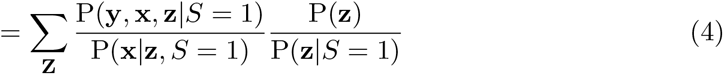

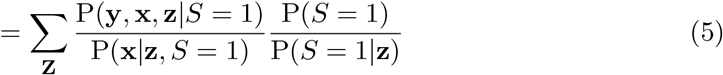

In the final expression, P(**y, x, z**|*S* = 1) is the joint distribution of outcome *Y*, exposure *X*, and auxiliary set **Z** observed under selection bias and P(**x**|**z**, *S* = 1) is analogous to a propensity score (CITE) under selection bias. The P(*S* = 1)*/*P(*S* = 1|**z**) term is the inverse probability of selection, or the weight used in IPW adjustment.

Methods for adjusting for selection bias all seek to identify such a set **Z** and estimate the inverse probability of selection weight, *w* = P(*S* = 1)*/*P(*S* = 1|**z**). They are differentiated by how **Z** is selected, and how *w* is estimated.

We evaluate the following six methods for adjusting for selection bias:

- Post-stratification
- Raking
- Calibration
- Raking with LASSO variable selection
- Logistic regression for estimating response propensity
- BART for estimating response propensity and raking

The first three methods (post-stratification, raking, and calibration) are standard survey inverse probability weighting techniques (Deville and Sarndal, 1992; Deville et al., 1993). These three methods are all examples of generalized raking procedures, differentiated by the specific measure used to regularize the distance between prior and posterior weight values. Practically, post-stratification considers the full joint distribution of categorical **Z**, while raking estimates *w* by iterating through the marginal distributions of each element of **Z** until weights converge. Calibration extends raking to allow elements in **Z** to be population totals rather than exclusively discrete variables.

*Raking with LASSO variable selection* seeks to address the problem of selecting a sufficient auxiliary set **Z** by using regularized regression of each *S* and **Y** on **Z** to select a subset of all covariates available in the population to serve as **Z**. After **Z** is selected, standard raking is performed.

*Logistic regression for estimating response propensity* and *BART for estimating response propensity and raking* both attempt to estimate P(*S* = 1)*/*P(*S* = 1|**z**) directly, without the constraint that the weighted distribution of **Z** must match the population distribution. *BART for estimating response propensity and raking* uses a Bayesian Additive Regression Tree (instead of simple logistic regression), which may better account for interactions between elements of **Z**, and has the additional step of standard raking using a subset of **Z** with the highest variable importance to ensure that the marginal distributions of key elements of **Z** match that of the population.

More details on the methods and their implementation can be found in Appendix B.

### 3.2 Simulation studies

We conduct simulation studies using UKB data to evaluate the relative performance of the six adjustment methods. In the simulation, we select random subsamples of various sizes from the 20,827 UK Biobank imaging subjects, and use adjustment procedures to estimate known quantities of the UK Biobank imaging population from the biased samples. First, we generate a missingness mechanism based on covariate data **Z** from the UK Biobank, and use it assign each subject *j* = (1, …, *N* = 20827) a probability *p*_*j*_ that they are observed (see Section C.1 for details on how the probability of missingness is generated).

On each iteration of the simulation, we randomly select a sample of a fixed size *n*_*obs*_ with probability proportional to *p*_*j*_. Then, we adjust that sample using each of the methods being considered. We perform this simulation 7 times, once for each *n*_*obs*_ ∈ *N* *(0.01, 0.02, 0.04, 0.05, 0.075, 0.1, 0.25). The full algorithm is described in 1.

Once we have generated weights for each sample, we calculate weighted estimates of the following quantities:

- **Brain volume**: total brain volume (gray and white matter), normalized for head size, measured in mm^3^ by T1 structural MRI. Brain volumes range from 1,151,700mm^3^ to 1,793,910mm^3^ with a mean of 1,502,37mm^3^,
- **Association between brain volume and age**: *β*_age_ from the weighted linear regression *Y*_brain volume_ = *β*_0_ + *β*_age_*Z*_age_ + *ϵ*

The simulation can be summarized as follows:

#### Algorithm 1. Simulation 1

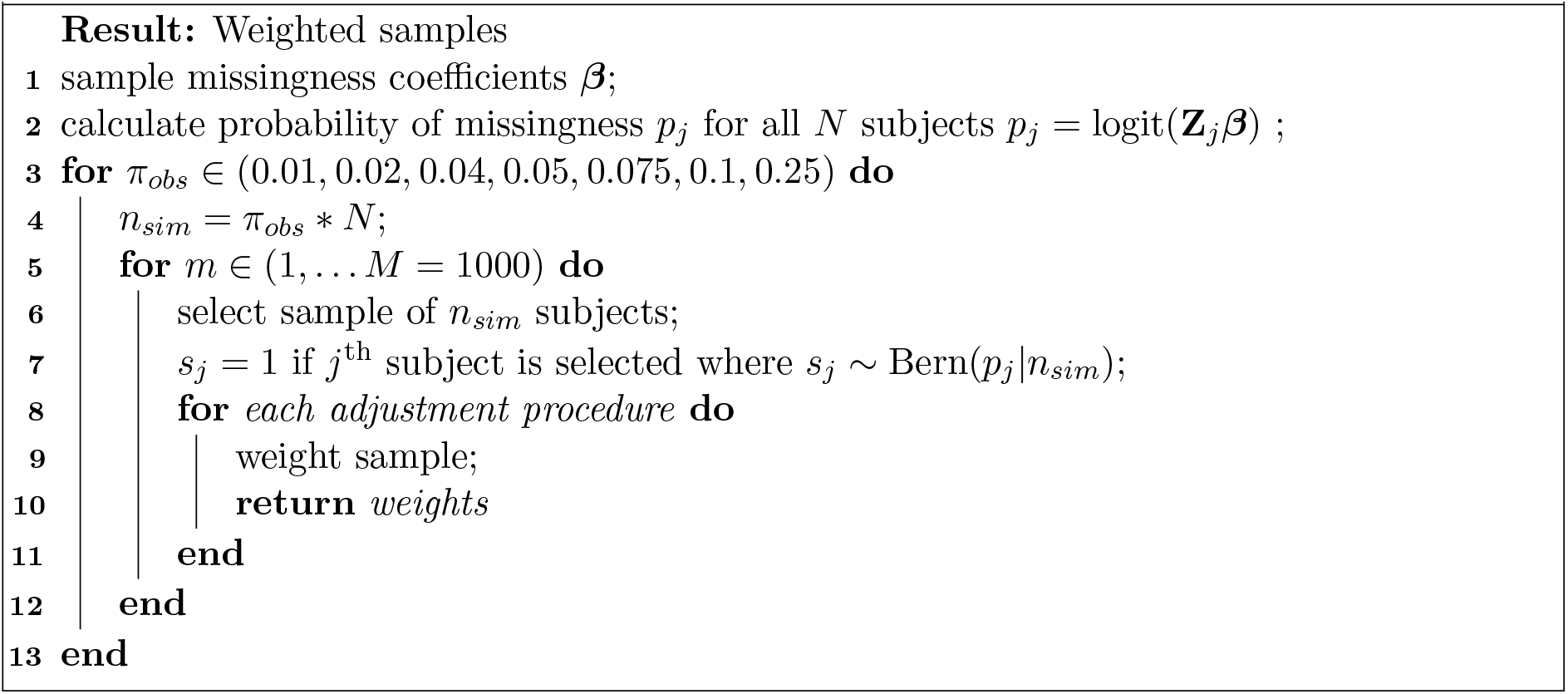

We will calculate the MSE for the weighted estimates produced by each method, as well as the design effect (Kish, 1992) in order to assess the “cost” (in terms of increased variance) of the reduction in bias from adjustment. We define one additional evaluation metric, **distribution bias**, as the sum of the squared distances between the weighted marginal distributions and target marginal distributions across variables used in adjustment. This metric will allow us to assess how well each adjustment method produces weighted marginal distributions of auxiliary variables that match those of the population. More details on the calculation can be found in Appendix C.2.

### 3.3 Application to the UK Biobank

After exploring the weighting methods in simulation studies, we apply them to the full UKB imaging cohort. We use each method to adjust the UKB imaging cohort to match the population distributions defined by the HSE. We use only demographic variables for adjustment, and assess the impact of adjustment on key heath outcomes. For health outcomes available in the HSE, we compare weighted UKB estimates to those from the HSE to assess the impact of weighting.

## 4 Results

### 4.1 Bias in the Neurological Imaging Cohort

Table 1 shows compares the UK Biobank imaging cohort to the 2016 HSE nurse interview sample. The table gives population counts (weighted in the case of the HSE) and the distribution of each study across levels of demographic variables.

**Table 1:**
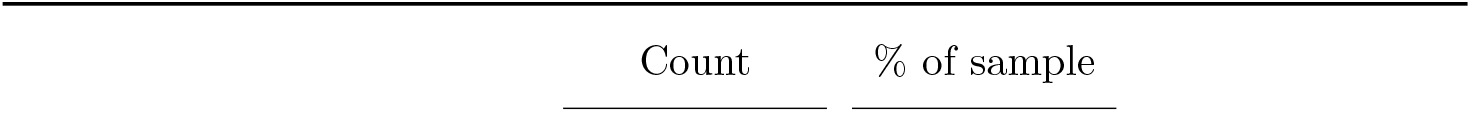
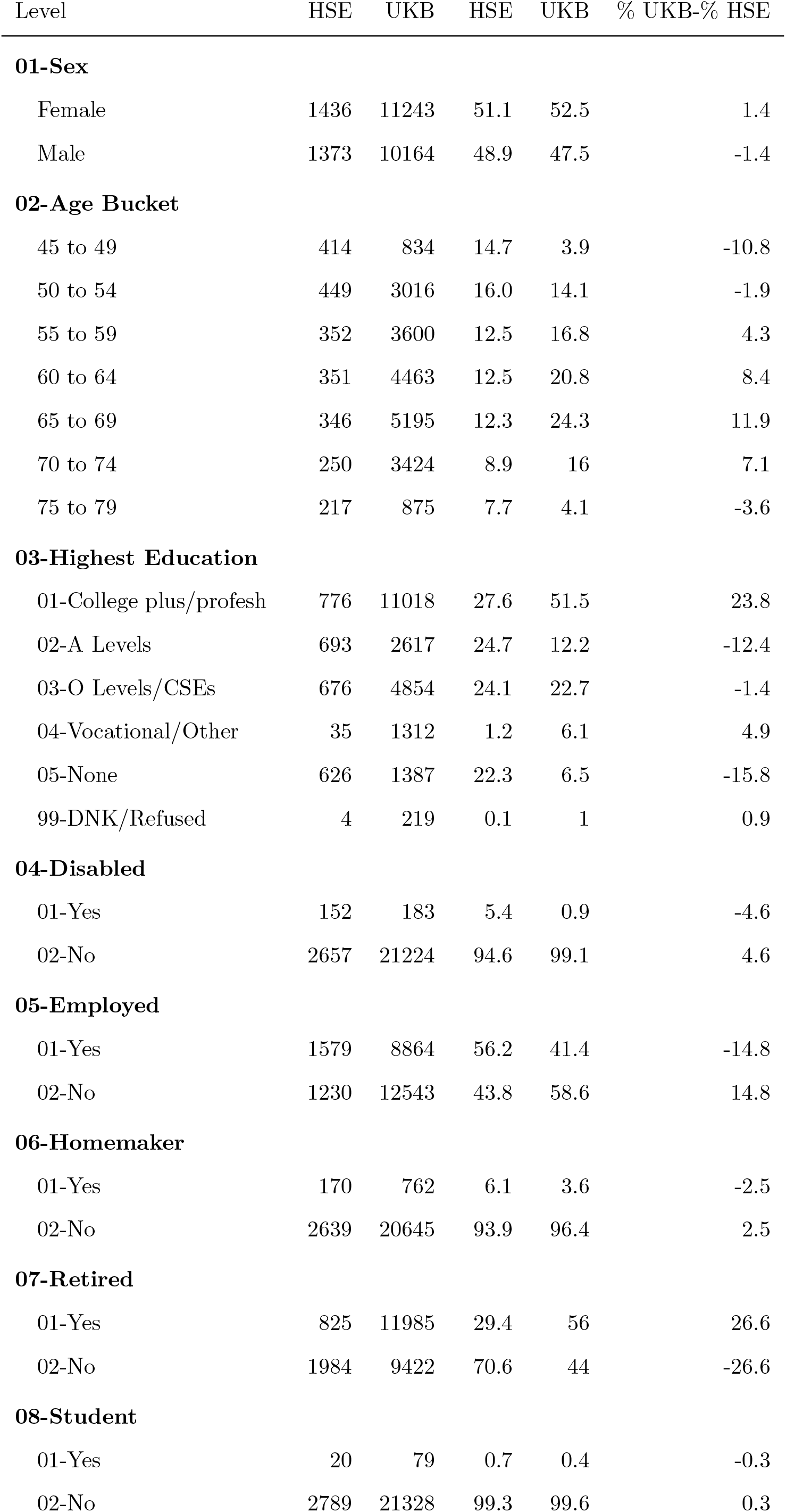
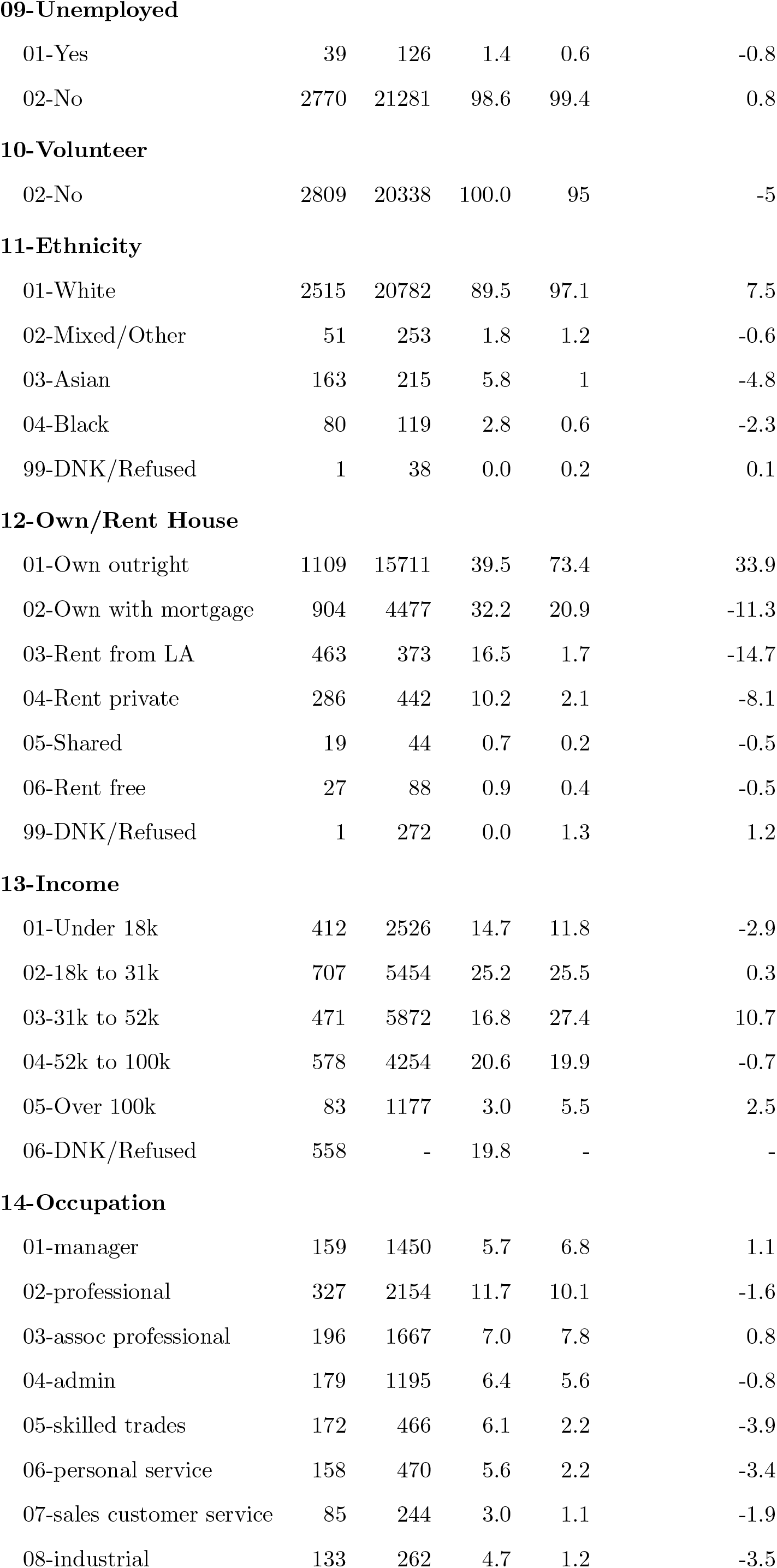
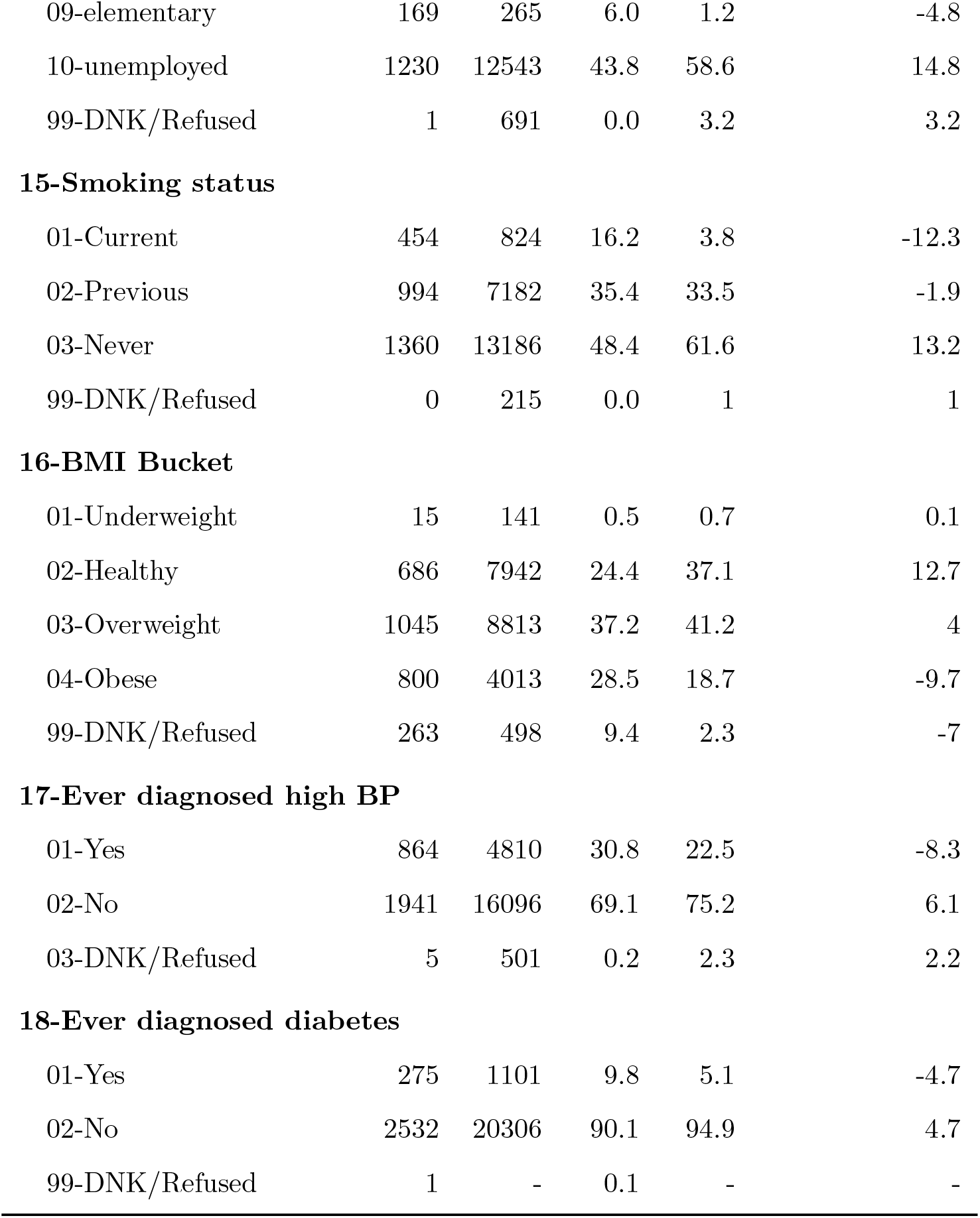
Selection bias in the UK Biobank imaging cohort relative to the 2016 HSE nurse interview subsample.

| Level | HSE | UKB | HSE | UKB | % UKB-% HSE |
| --- | --- | --- | --- | --- | --- |
| **01-Sex** |  |  |  |  |  |
| Female | 1436 | 11243 | 51.1 | 52.5 | 1.4 |
| Male | 1373 | 10164 | 48.9 | 47.5 | -1.4 |
| **02-Age Bucket** |  |  |  |  |  |
| 45 to 49 | 414 | 834 | 14.7 | 3.9 | -10.8 |
| 50 to 54 | 449 | 3016 | 16.0 | 14.1 | -1.9 |
| 55 to 59 | 352 | 3600 | 12.5 | 16.8 | 4.3 |
| 60 to 64 | 351 | 4463 | 12.5 | 20.8 | 8.4 |
| 65 to 69 | 346 | 5195 | 12.3 | 24.3 | 11.9 |
| 70 to 74 | 250 | 3424 | 8.9 | 16 | 7.1 |
| 75 to 79 | 217 | 875 | 7.7 | 4.1 | -3.6 |
| **03-Highest Education** |  |  |  |  |  |
| 01-College plus/profesh | 776 | 11018 | 27.6 | 51.5 | 23.8 |
| 02-A Levels | 693 | 2617 | 24.7 | 12.2 | -12.4 |
| 03-O Levels/CSEs | 676 | 4854 | 24.1 | 22.7 | -1.4 |
| 04-Vocational/Other | 35 | 1312 | 1.2 | 6.1 | 4.9 |
| 05-None | 626 | 1387 | 22.3 | 6.5 | -15.8 |
| 99-DNK/Refused | 4 | 219 | 0.1 | 1 | 0.9 |
| **04-Disabled** |  |  |  |  |  |
| 01-Yes | 152 | 183 | 5.4 | 0.9 | -4.6 |
| 02-No | 2657 | 21224 | 94.6 | 99.1 | 4.6 |
| **05-Employed** |  |  |  |  |  |
| 01-Yes | 1579 | 8864 | 56.2 | 41.4 | -14.8 |
| 02-No | 1230 | 12543 | 43.8 | 58.6 | 14.8 |
| **06-Homemaker** |  |  |  |  |  |
| 01-Yes | 170 | 762 | 6.1 | 3.6 | -2.5 |
| 02-No | 2639 | 20645 | 93.9 | 96.4 | 2.5 |
| **07-Retired** |  |  |  |  |  |
| 01-Yes | 825 | 11985 | 29.4 | 56 | 26.6 |
| 02-No | 1984 | 9422 | 70.6 | 44 | -26.6 |
| **08-Student** |  |  |  |  |  |
| 01-Yes | 20 | 79 | 0.7 | 0.4 | -0.3 |
| 02-No | 2789 | 21328 | 99.3 | 99.6 | 0.3 |
| **09-Unemployed** |  |  |  |  |  |
| 01-Yes | 39 | 126 | 1.4 | 0.6 | -0.8 |
| 02-No | 2770 | 21281 | 98.6 | 99.4 | 0.8 |
| **10-Volunteer** |  |  |  |  |  |
| 02-No | 2809 | 20338 | 100.0 | 95 | -5 |
| **11-Ethnicity** |  |  |  |  |  |
| 01-White | 2515 | 20782 | 89.5 | 97.1 | 7.5 |
| 02-Mixed/Other | 51 | 253 | 1.8 | 1.2 | -0.6 |
| 03-Asian | 163 | 215 | 5.8 | 1 | -4.8 |
| 04-Black | 80 | 119 | 2.8 | 0.6 | -2.3 |
| 99-DNK/Refused | 1 | 38 | 0.0 | 0.2 | 0.1 |
| **12-Own/Rent House** |  |  |  |  |  |
| 01-Own outright | 1109 | 15711 | 39.5 | 73.4 | 33.9 |
| 02-Own with mortgage | 904 | 4477 | 32.2 | 20.9 | -11.3 |
| 03-Rent from LA | 463 | 373 | 16.5 | 1.7 | -14.7 |
| 04-Rent private | 286 | 442 | 10.2 | 2.1 | -8.1 |
| 05-Shared | 19 | 44 | 0.7 | 0.2 | -0.5 |
| 06-Rent free | 27 | 88 | 0.9 | 0.4 | -0.5 |
| 99-DNK/Refused | 1 | 272 | 0.0 | 1.3 | 1.2 |
| **13-Income** |  |  |  |  |  |
| 01-Under 18k | 412 | 2526 | 14.7 | 11.8 | -2.9 |
| 02-18k to 31k | 707 | 5454 | 25.2 | 25.5 | 0.3 |
| 03-31k to 52k | 471 | 5872 | 16.8 | 27.4 | 10.7 |
| 04-52k to 100k | 578 | 4254 | 20.6 | 19.9 | -0.7 |
| 05-Over 100k | 83 | 1177 | 3.0 | 5.5 | 2.5 |
| 06-DNK/Refused | 558 | - | 19.8 | - | - |
| **14-Occupation** |  |  |  |  |  |
| 01-manager | 159 | 1450 | 5.7 | 6.8 | 1.1 |
| 02-professional | 327 | 2154 | 11.7 | 10.1 | -1.6 |
| 03-assoc professional | 196 | 1667 | 7.0 | 7.8 | 0.8 |
| 04-admin | 179 | 1195 | 6.4 | 5.6 | -0.8 |
| 05-skilled trades | 172 | 466 | 6.1 | 2.2 | -3.9 |
| 06-personal service | 158 | 470 | 5.6 | 2.2 | -3.4 |
| 07-sales customer service | 85 | 244 | 3.0 | 1.1 | -1.9 |
| 08-industrial | 133 | 262 | 4.7 | 1.2 | -3.5 |
| 09-elementary | 169 | 265 | 6.0 | 1.2 | -4.8 |
| 10-unemployed | 1230 | 12543 | 43.8 | 58.6 | 14.8 |
| 99-DNK/Refused | 1 | 691 | 0.0 | 3.2 | 3.2 |
| **15-Smoking status** |  |  |  |  |  |
| 01-Current | 454 | 824 | 16.2 | 3.8 | -12.3 |
| 02-Previous | 994 | 7182 | 35.4 | 33.5 | -1.9 |
| 03-Never | 1360 | 13186 | 48.4 | 61.6 | 13.2 |
| 99-DNK/Refused | 0 | 215 | 0.0 | 1 | 1 |
| **16-BMI Bucket** |  |  |  |  |  |
| 01-Underweight | 15 | 141 | 0.5 | 0.7 | 0.1 |
| 02-Healthy | 686 | 7942 | 24.4 | 37.1 | 12.7 |
| 03-Overweight | 1045 | 8813 | 37.2 | 41.2 | 4 |
| 04-Obese | 800 | 4013 | 28.5 | 18.7 | -9.7 |
| 99-DNK/Refused | 263 | 498 | 9.4 | 2.3 | -7 |
| **17-Ever diagnosed high BP** |  |  |  |  |  |
| 01-Yes | 864 | 4810 | 30.8 | 22.5 | -8.3 |
| 02-No | 1941 | 16096 | 69.1 | 75.2 | 6.1 |
| 03-DNK/Refused | 5 | 501 | 0.2 | 2.3 | 2.2 |
| **18-Ever diagnosed diabetes** |  |  |  |  |  |
| 01-Yes | 275 | 1101 | 9.8 | 5.1 | -4.7 |
| 02-No | 2532 | 20306 | 90.1 | 94.9 | 4.7 |
| 99-DNK/Refused | 1 | - | 0.1 | - | - |

#### Sociodemographic factors

The UK Biobank imaging cohort is older than the HSE (44.4% and 28.9% aged 65 or older, respectively), more educated (51.5% v. 27.6%), more white (97.1% v. 89.5%) and more likely to be retired (56% v. 39.4%) or to own a home (73.4% v. 39.5%). Notably, the cohort does not have a gender bias (52% women compared to 51.5% in the HSE), despite the higher participation rates among women found by Fry et al. (2017).

#### Health characteristics

The UK Biobank imaging cohort is healthier than the general population. Only 3.8% are current smokers, compared to 16.2% of the general population. The cohort is less likely to be obese (18.7%) than the HSE (28.5%), to have ever been diagnosed with high blood pressure (22.5% v. 30.8%) or to have ever been diagnosed with diabetes (5.1% v. 9.8%).

### 4.2 Simulation results

Results from simulation studies are arranged by outcome of interest: total brain volume, association between total brain volume and ApoE, and population composition.

#### 4.2.1 Total brain volume

Figure 1 shows the distribution of mean total brain volume in our random subsamples, All samples have mean total brain volumes far below the true poplation value, showing that we successfully induced selection bias.

**Figure 1:**
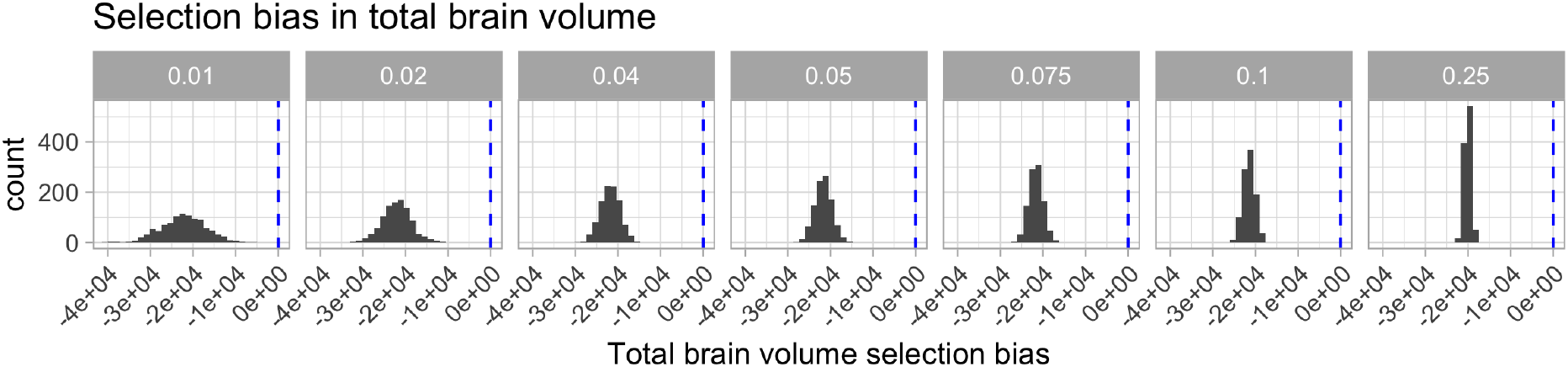
Histogram of mean total brain volume in simulated samples. The blue dotted line represents the true population mean total brain volume.

##### Bias

Figure 2 shows the bias in estimated total brain volume remaining after adjustment across weighting methods and sample sizes. At the smallest sample size (208), the BART method produces the best estimate of population total brain volume, followed by stratification and raking. BART produces unbiased estimates at larger sample sizes, until a slight dip when the sample is a full 25% of the population. At larger sample sizes, 833 and above, post-stratification performs at least as well as BART. The magnitude of the bias in the logit-weighted estimator decreases consistently as sample size increases. Calibration, lasso and raking estimators are not unbiased at any sample size.

**Figure 2:**
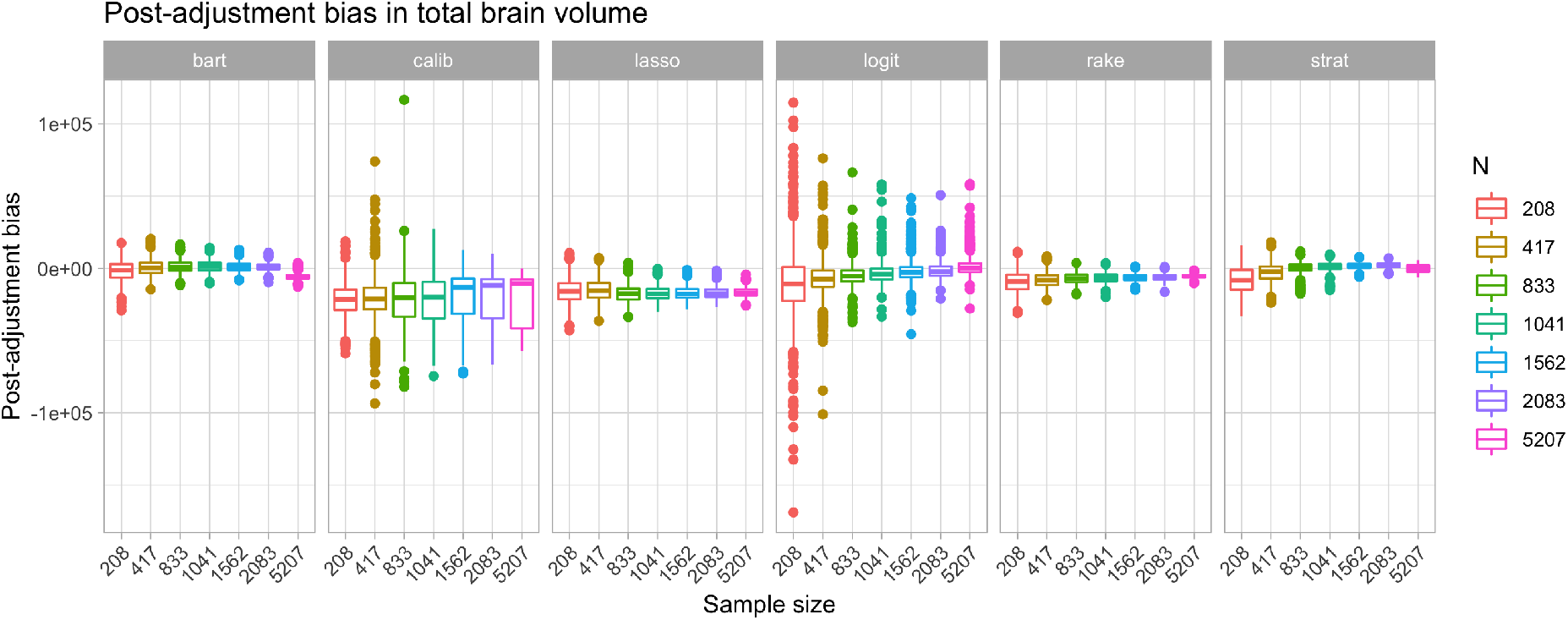
Error by method and sample size in simulation studies

##### Design effect

Figure 3 shows the design effect of each method across sample sizes. The calibration and logit weights have considerably larger design effects, and design effects that are themselves highly variable. The LASSO method has the smallest design effect across all sample sizes. BART and raking have similarly small design effects, consistent across sample sizes, while post-stratification has small design effects that increase slightly with sample size.

**Figure 3:**
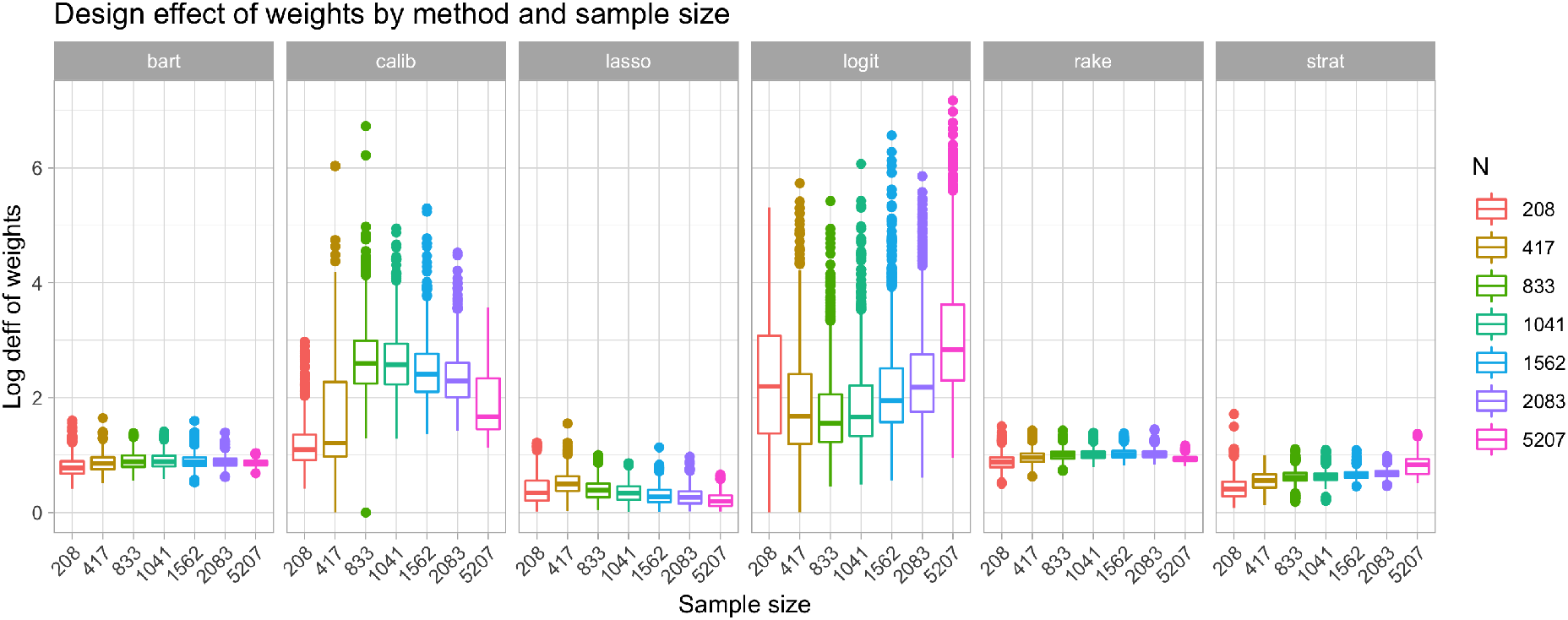
Log design effect by method and sample size in simulation studies

##### MSE

Figure 4a shows the log MSE of total brain volume estimators as a function of the proportion of the population sampled. The BART estimator has the lowest MSE for 1% and 2% of the population sampled, but is surpassed by post-stratification at larger sample sizes.

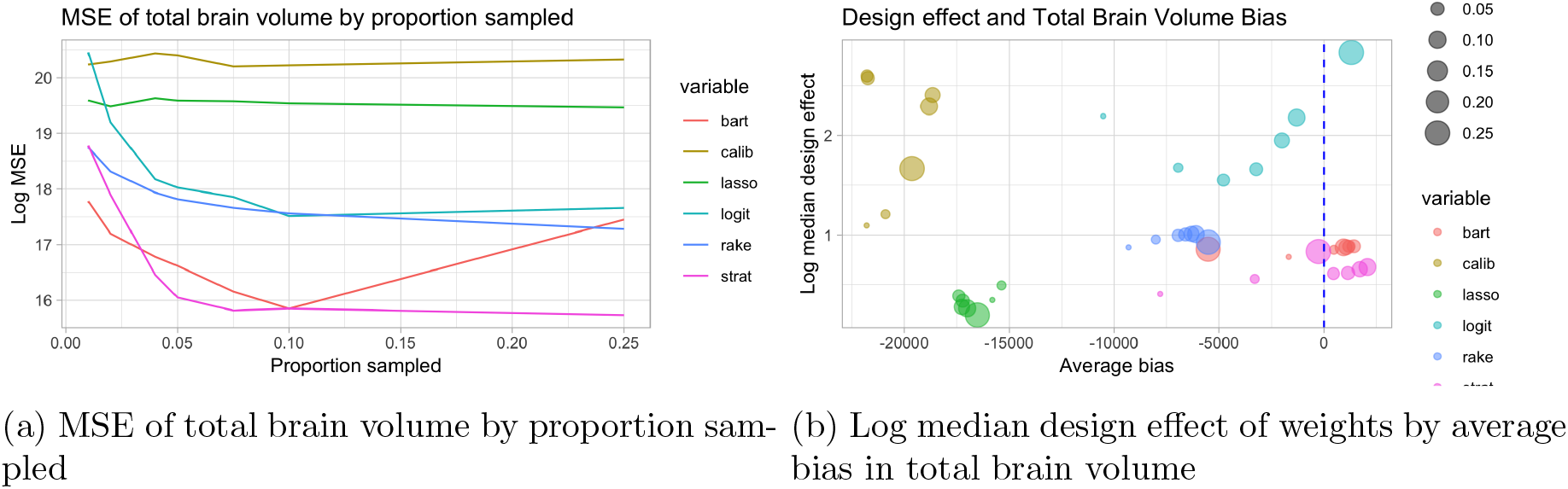

The logit and raking estimators perform similarly, with a sharp drop in MSE as the proportion sampled increases until 5%, then a steady decrease. Calibration and the LASSO have consistent, high MSE compared to the other methods.

##### Bias and design effect

The ideal estimator will eliminate bias without a introducing a large design effect. Figure 4b shows the relationship between the log median bias and the log median design effect across methods (shown in color) and proportion sampled (size of each point). We use log medians due to the large skew in the distributions of each variable. The LASSO estimator has the smallest design effect, but hardly eliminates bias. Conversely, the logit effectively eliminates bias as proportion sampled increases, but at the cost of a large design effect. Calibration is clearly the worst performing, as it fails to reduce bias and also has a large design effect. BART and post-stratification both effectively eliminate bias without a large design effect. BART has a small advantage at the smaller sample sizes, but post-stratification quickly catches up.

##### Subgroup estimation

Trends from the metrics discussed thus far persist at the subgroup level. Figure 5 shows log MSE of total brain volume within key subgroups as a function of the true size of the subgroup in the population. There is no clear difference in performance between methods in the smallest subsets, however BART has the lowest MSE for larger subgroups in the two smallest samples, slightly outperforming raking and post-stratification. Post-stratification outperforms other methods in larger subgroups as sampled proportion increases. Calibration consistently has the highest MSE, with the LASSO not far behind.

**Figure 5:**
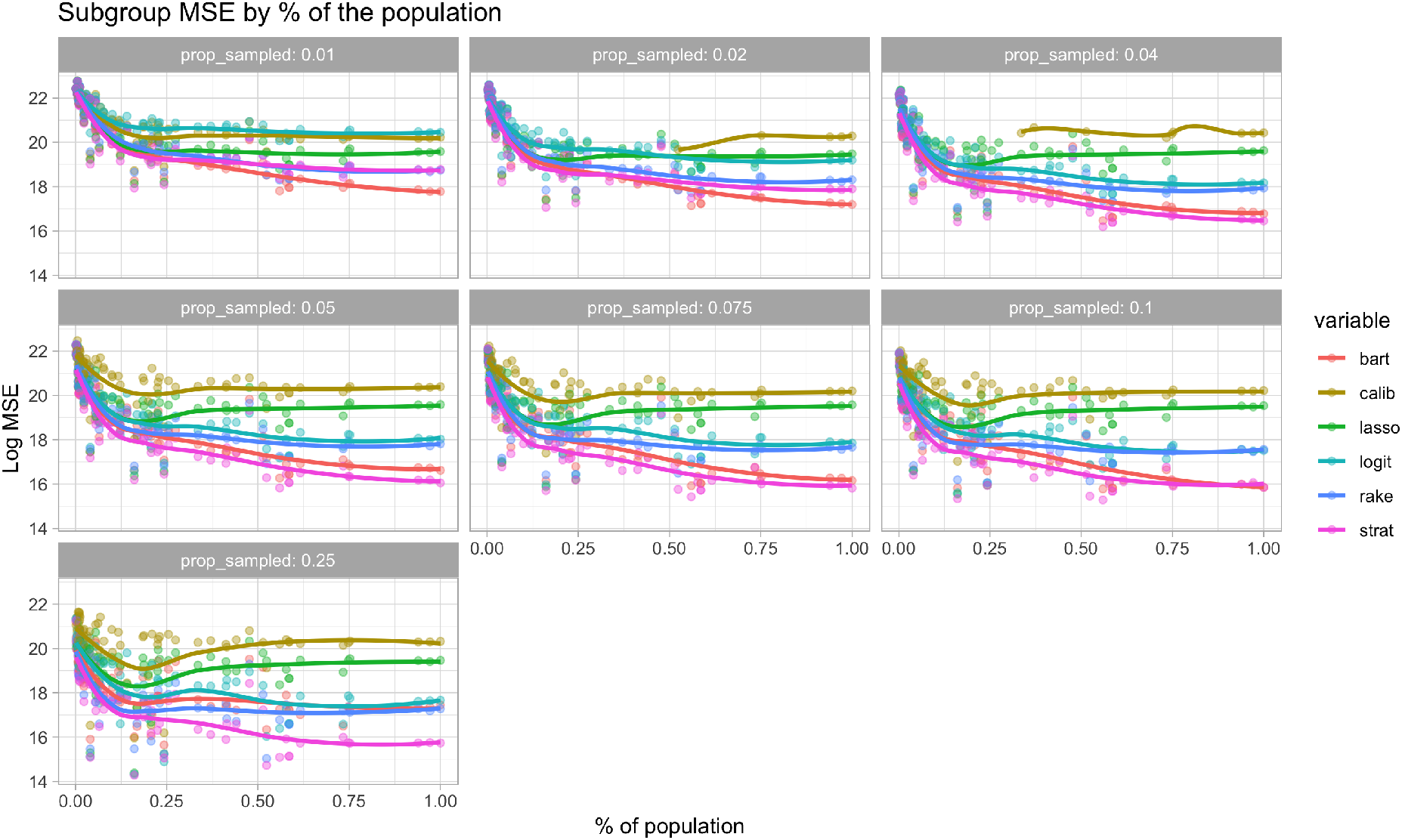
Points represent the log MSE of total brain volume calculated for demographic subgroups, by the true proportion of the population made up by that subgroup. Lines are smoothed estimates of the association between proportion of the population and MSE.

#### 4.2.2 Age and total brain volume

For each sample, we regress total brain volume on age, once in a simple linear regression, and once in a weighted linear regression for each adjustment method. We record the estimated values of the intercept and age coefficient. Figure 6 shows the simulated selection bias in the age coefficient from these regressions, estimated with the unweighted linear regression. The histograms are distributions of the age coefficient, and the blue dotted line represents population truth. We can see that there is a small amount of selection bias in the unadjusted age coefficients. Figure 7 shows the bias in the age coefficient remaining after adjustment procedures were applied. While it appears that all procedures are relatively unbiased, this is more a function of the small amount of selection bias introduced (as a proportion of the total variation in the age coefficient) rather than performance of the methods. The logit approach seems to produce the only truly unbiased estimator of the age coefficient, however we can see in Figure 8 that the MSE oof the logit method is still quite high. Though BART and post-stratification don’t seem to completely eliminate the selection bias in the estimate of the association between total brain volume and age, they do so without introducting a large amount of variance, so on the whole seem to perform slightly better than other methods.

**Figure 6:**
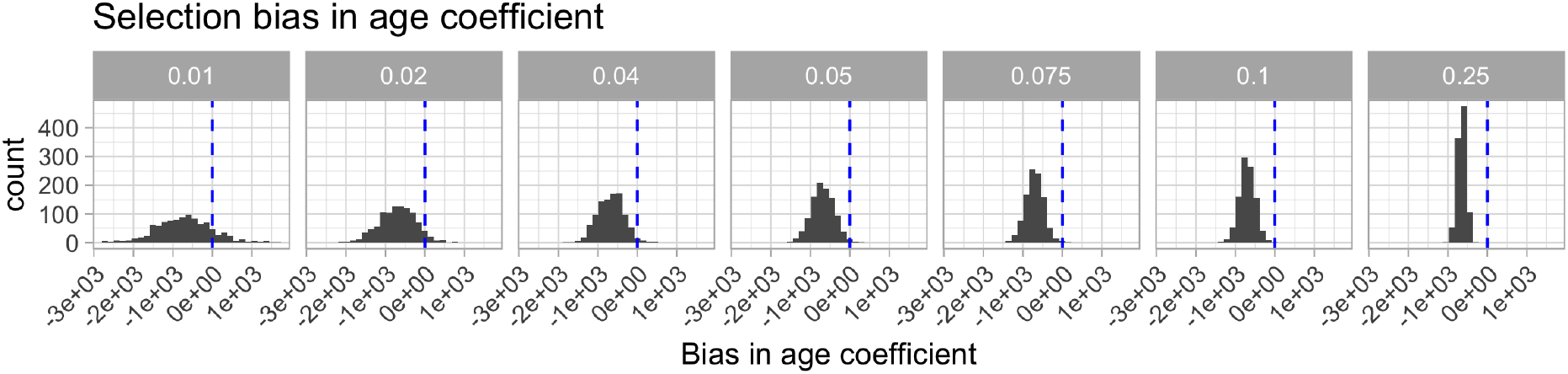
The distribution of estimates of the coefficient for age in a linear regression of total brain volume on age. Regressions were estimated using unadjusted (selection-biased) sample data. The blue lines represent the true population value of the age coefficient.

**Figure 7:**
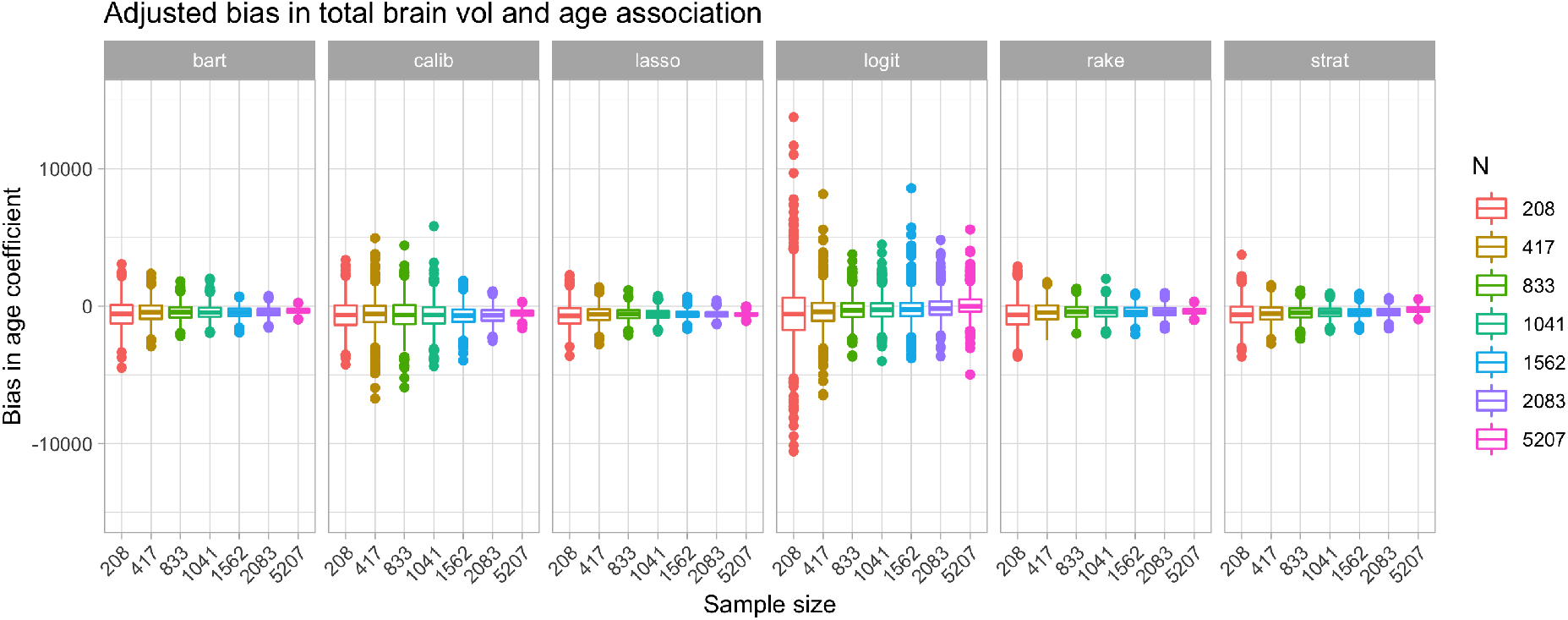
Bias in weighted estimate of age coefficient in regression predicting total brain volume.

**Figure 8:**
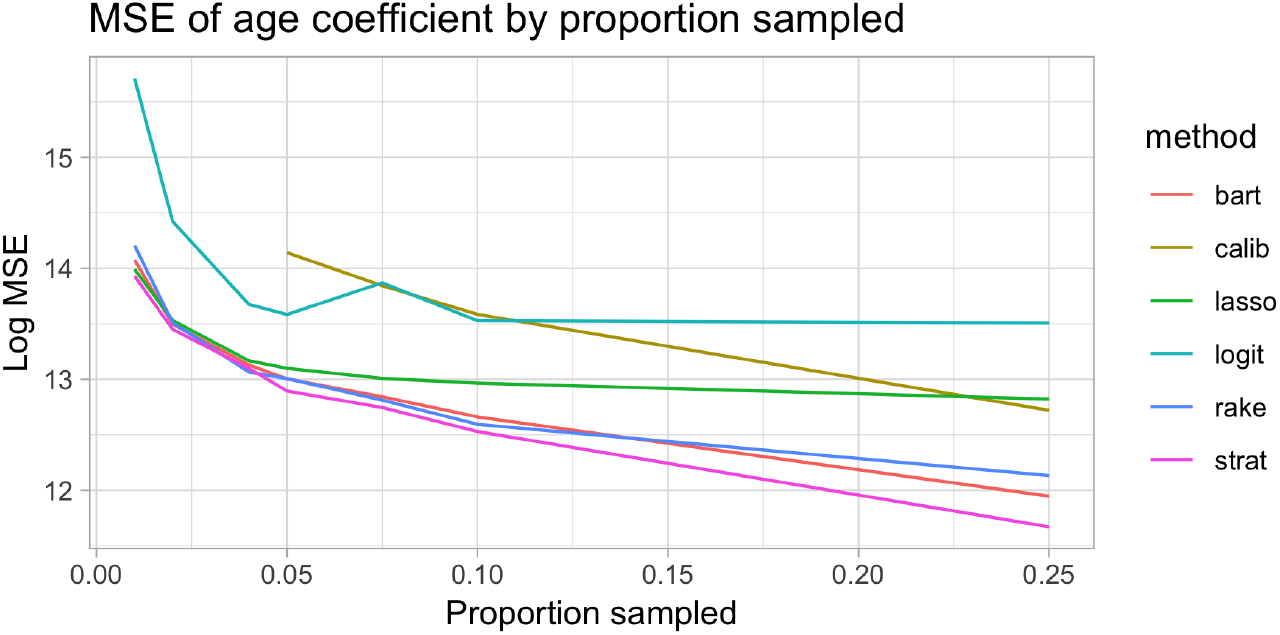
Log MSE as a function of proportion sampled. in larger sample sizes than raking or logit.

#### 4.2.3 Population composition

The last metric by which we evaluate adjustment methods is distribution bias (DB). Figure 9 shows the distribution of DB across simulations for each adjustment method by sample size. Raking has the smallest DB across all sample sizes considered, and the DB decreased as sample size grew. DB for the logit method was the most variable, but, on average, decreased the most drastically with increases in sample size. DB for both post-stratification and BART was less variable across simulation iterations, and consistent across sample sizes, but higher on average

**Figure 9:**
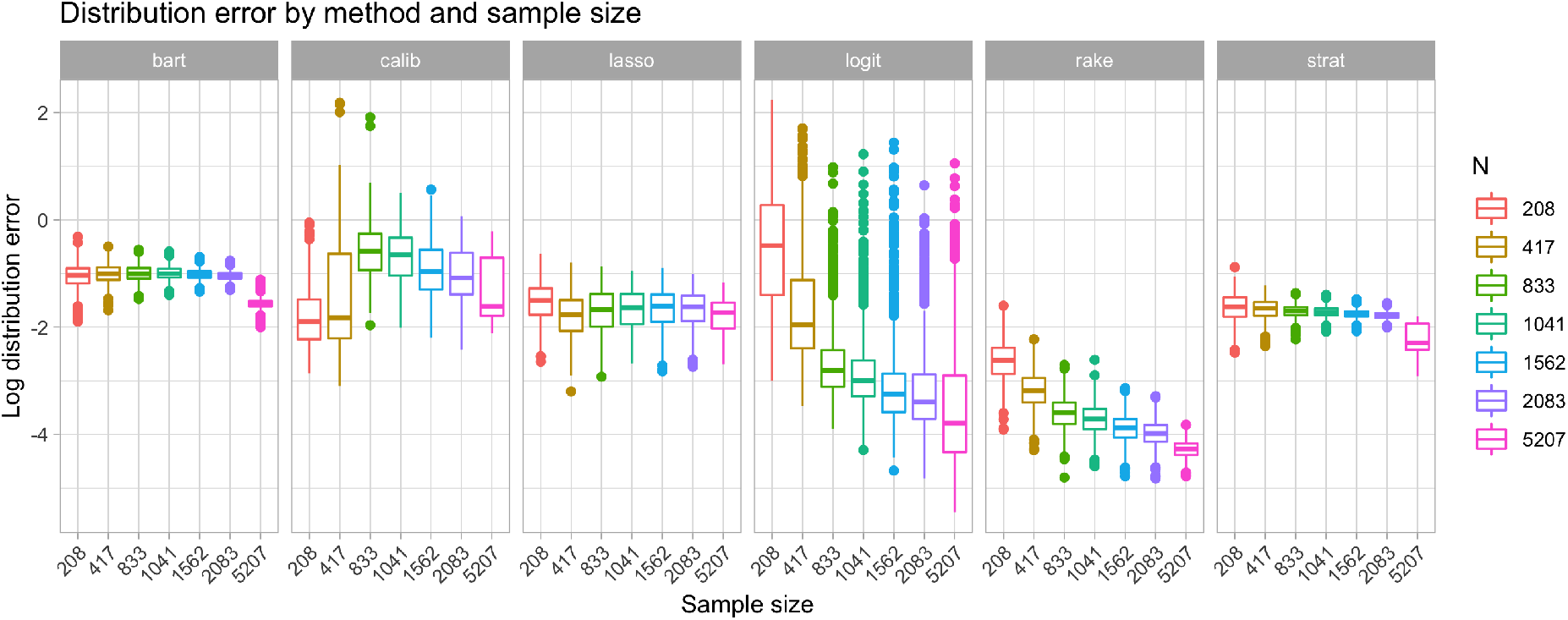
Log distribution bias by method adjustment method and sample size.

### 4.3 Application to the UK Biobank

Table 2 gives adjusted estimates of prevalence for selected health outcomes from the UK Biobank imaging cohort, related to unadjusted UK Biobank data and population estimates from the HSE. Weighting generally seems to improve prevalance estimates for most health outcomes.

**Table 2:**
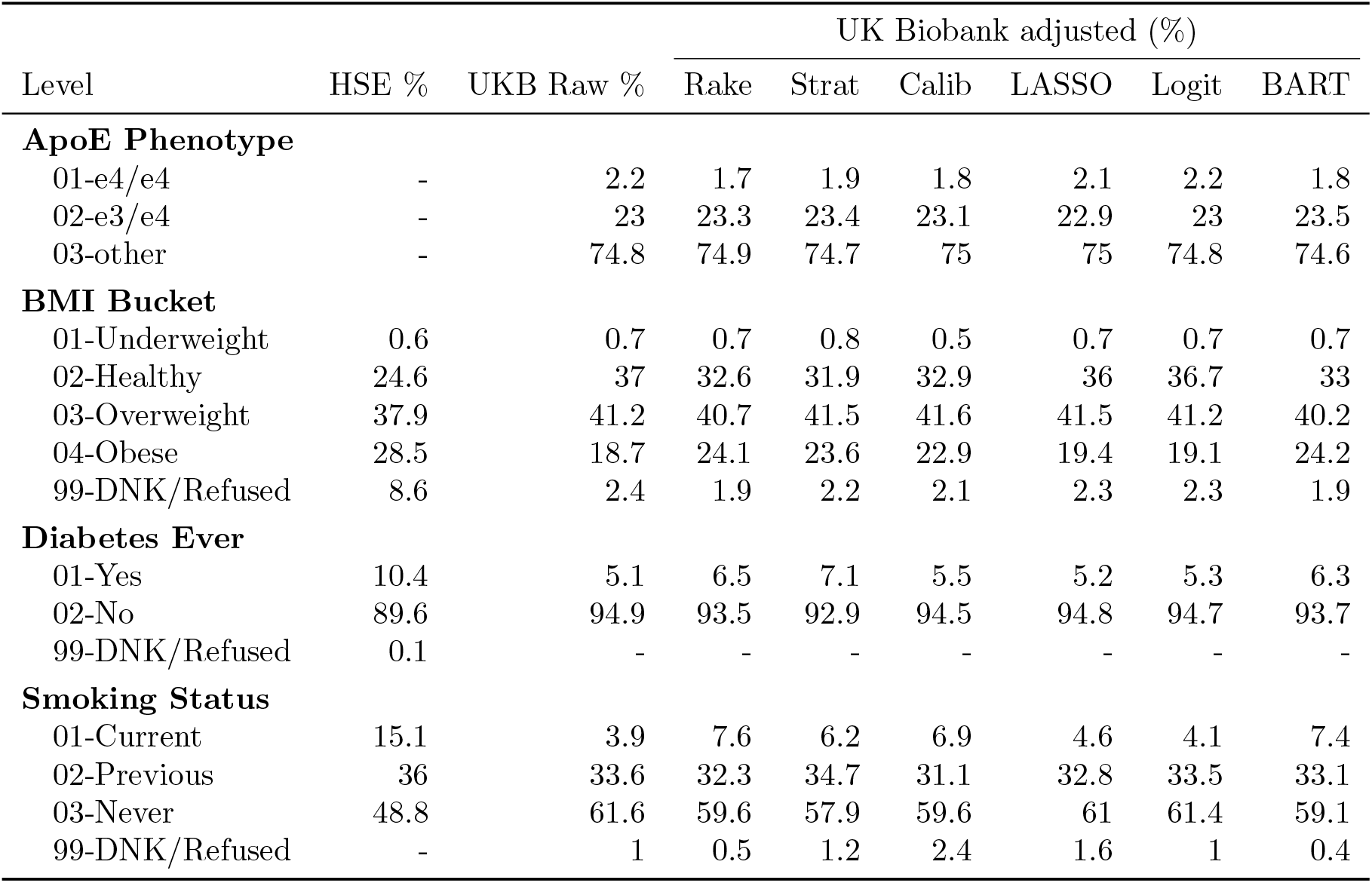
Weighted estimates of prevalence health outcomes from the UK Biobank imaging cohort data, compared to our target population, the HSE.

| Level | HSE % | UKB Raw % | UK Biobank adjusted (%) |  |  |  |  |  |
| --- | --- | --- | --- | --- | --- | --- | --- | --- |
|  |  |  | Rake | Strat | Calib | LASSO | Logit | BART |
| ApoE Phenotype |  |  |  |  |  |  |  |  |
| 01-e4/e4 | - | 2.2 | 1.7 | 1.9 | 1.8 | 2.1 | 2.2 | 1.8 |
| 02-e3/e4 | - | 23 | 23.3 | 23.4 | 23.1 | 22.9 | 23 | 23.5 |
| 03-other | - | 74.8 | 74.9 | 74.7 | 75 | 75 | 74.8 | 74.6 |
| BMI Bucket |  |  |  |  |  |  |  |  |
| 01-Underweight | 0.6 | 0.7 | 0.7 | 0.8 | 0.5 | 0.7 | 0.7 | 0.7 |
| 02-Healthy | 24.6 | 37 | 32.6 | 31.9 | 32.9 | 36 | 36.7 | 33 |
| 03-Overweight | 37.9 | 41.2 | 40.7 | 41.5 | 41.6 | 41.5 | 41.2 | 40.2 |
| 04-Obese | 28.5 | 18.7 | 24.1 | 23.6 | 22.9 | 19.4 | 19.1 | 24.2 |
| 99-DNK/Refused | 8.6 | 2.4 | 1.9 | 2.2 | 2.1 | 2.3 | 2.3 | 1.9 |
| Diabetes Ever |  |  |  |  |  |  |  |  |
| 01-Yes | 10.4 | 5.1 | 6.5 | 7.1 | 5.5 | 5.2 | 5.3 | 6.3 |
| 02-No | 89.6 | 94.9 | 93.5 | 92.9 | 94.5 | 94.8 | 94.7 | 93.7 |
| 99-DNK/Refused | 0.1 | - | - | - | - | - | - | - |
| Smoking Status |  |  |  |  |  |  |  |  |
| 01-Current | 15.1 | 3.9 | 7.6 | 6.2 | 6.9 | 4.6 | 4.1 | 7.4 |
| 02-Previous | 36 | 33.6 | 32.3 | 34.7 | 31.1 | 32.8 | 33.5 | 33.1 |
| 03-Never | 48.8 | 61.6 | 59.6 | 57.9 | 59.6 | 61 | 61.4 | 59.1 |
| 99-DNK/Refused | - | 1 | 0.5 | 1.2 | 2.4 | 1.6 | 1 | 0.4 |

Take, for example, the proportion of the population estimated to be current smokers. The HSE estimates that 15.1% of the population smokes, while only 3.9% of the subjects in the UK Biobank imaging cohort report being current smokers. All methods of adjustment improve this estimatet. BART and raking estimate that 7.4% and 7.6% of the population currently smokes, while the lowest estimate is from the LASSO which estimates that 4.6% of the population smokes.

Similarly, for obesity, BART estimates that 24.2% of the population is obese, compared to 28.5% in the HSE, while the unadjusted estimate is only 18.7%. Other methods improve on the unadjusted estimate, however none so much as BART. Stratification also seems to drastically improve estimates of these health quantities, while other methods, like the LASSO and logit, seem to lag behind. It is important to note that while these estimators improve on the unadjusted estimator, even the best-performing are not able to completely eliminate selection bias.

Table 3 gives weighted estimates of total brain volume relative to the unweighted estimate from the UK Biobank imaging cohort. The weighted estimates have hardly changed from the unweighted estimate (the largest difference is from BART, which changes only by about 1% of the unweighted total brain volume estimate).

**Table 3:**
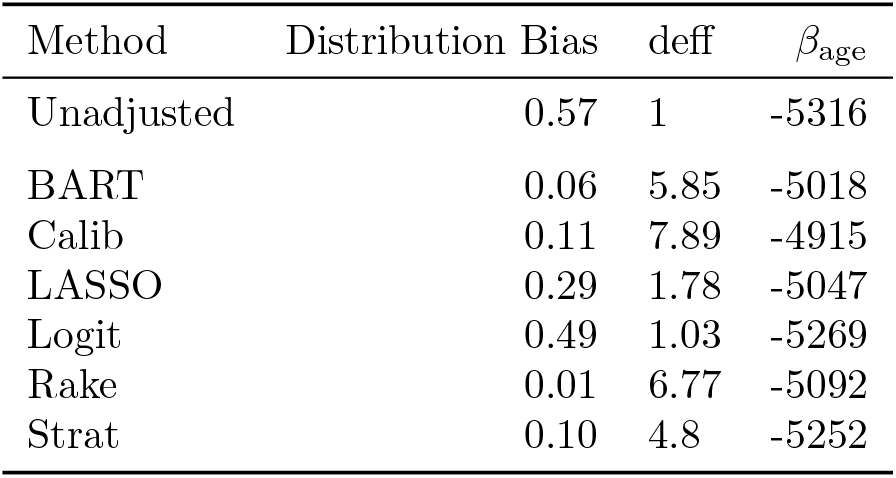
Adjusted estimates of total brain volume from the UK Biobank imaging cohort

| Method | Distribution Bias | deff | $\beta_{\text{age}}$ |
| --- | --- | --- | --- |
| Unadjusted | 0.57 | 1 | -5316 |
| BART | 0.06 | 5.85 | -5018 |
| Calib | 0.11 | 7.89 | -4915 |
| LASSO | 0.29 | 1.78 | -5047 |
| Logit | 0.49 | 1.03 | -5269 |
| Rake | 0.01 | 6.77 | -5092 |
| Strat | 0.10 | 4.8 | -5252 |

Table 4 shows the deisign effect, distribuion bias and estimated age coefficient for each method. The results are in-line with simulation studies. Raking is the best at matching population totals of auxiliary variables (as measuured by the lowest DB), with BART and poststratification performing almost as well. Calibration matching population totals reasonably well, but has the largest design effect. The logit approach does not reduce distribution bias from the unweighted estimate, which is consistent with the known shortcomings of the method. BART, as a regression-based approach, improves considerably on the logit baseline.

**Table 4:**
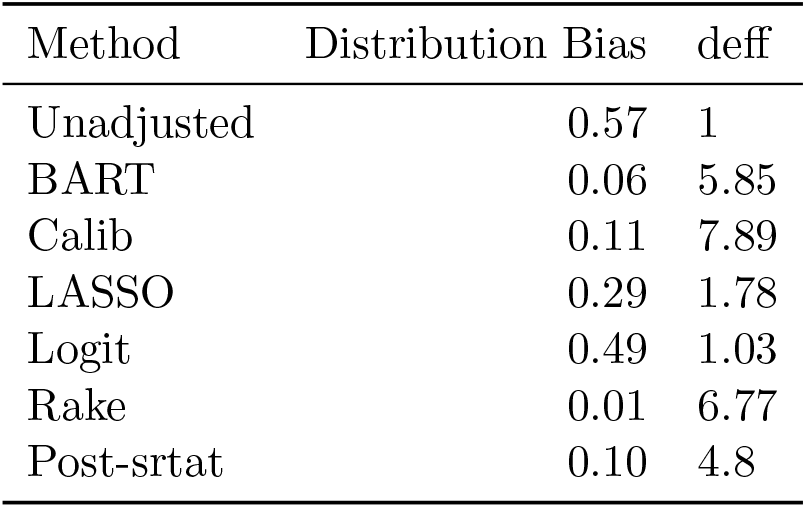
The distribution bias (DB) and design effect (deff) of each adjustment method relative to unadjusted estimates.

| Method | Distribution Bias | deff |
| --- | --- | --- |
| Unadjusted | 0.57 | 1 |
| BART | 0.06 | 5.85 |
| Calib | 0.11 | 7.89 |
| LASSO | 0.29 | 1.78 |
| Logit | 0.49 | 1.03 |
| Rake | 0.01 | 6.77 |
| Post-srtat | 0.10 | 4.8 |

Age coefficients are adjusted down from the unweighted estimator, but not to a significant degree. There is not much differentiation in estimated association between age and total brain volume across adjustment methods.

## 5 Discussion

The simulation studies revealed BART and post-stratification to be the most effective at reducing bias without introducing much additional variance. BART slightly outperformed poststratification in the smallest samples, likely the more realistic scenarios than observing 25% of the target population. BART, however, only did a mediocre job of matching sample marginal distributions to those of the population. This is likely due to the fact that raking variables were selected based on which were the most important predictors of selection, and not based on which represented the largest subgroups in the population. Altering this selection criterion could improve BART’s performance along this metric.

Calibration performed almost remarkably poorly, failing to reduce bias while drastically increasing variance compared to other methods. It performed far worse than raking, to which its closely related, except that calibration included constraints based on the continuous form of age, while raking relied on only the discrete specification of age. It is possible that since we only calculated performance of methods using categorical forms of variables, our assessment did not fully capture advantages of calibration. This could also be due to poor specification of algorithm parameters, and warrants further examination.

That LASSO performed poorly was also surprising, as it is simply raking with a variable selection step. It may be that the LASSO is not able to reliably select the most important variables for adjustment, or that our method of creating tiers of variables for sequential raking is not the optimal strategy for handling a large number of selected variables.

From an implementation standpoint, BART has two main advantages over other methods: variable selection is done automatically, and complex interactions are implicitly considered without the need to enumerate all of them. LASSO and logit include a mechanism for variable selection, but seem to lack the power that BART has to identify critical variables in small samples. They also both select interactions only from a set previously specified by the researcher, which limits the degree of interactions that can be considered before hitting computation time and memory errors. Post-stratification, in the way we have implemented it here, also considers interactions and has an automatic variable selection feature, likely why it performs similarly to BART. However, in small samples, there is a limit to the number of variables that can be used for post-stratification before strata become too small. BART has no such limitation, likely why it outperforms post-stratification in smaller samples. In large samples, post-stratification can consider high-degrees of interactions, and will dominate performance.

Logit adjustment is the only method without specific constraints of matching weighted sample marginal distributions to population distributions, however, in small samples had a DB on par with other methods, on average, and in larger samples had some of the lowest DB of any method. One caveat is that though the DB was low on average, it was highly variable.

It is important to note that most of the methods that we consider here require access not only to external population data, but specifically to individual-level population data, which is not always available. Our best-performing methods, BART and post-stratification, are not possible without access at least to joint distributions of auxiliary variables. Other methods, like raking and calibration, though they perform worse in this setting are still highly useful in other scenarios.

In the application of these methods to real UK Biobank data, we observed that they were able to improve estimates of prevalence of smoking, diabetes, obesity and high blood pressure relative to population estimates from the HSE. BART and post-stratification exhibited the largest improvements over the unweighted estimators, though were still unable to eliminate selection bias completely.

There are numerous caveats and limitations of the results presented here. First, the data we use as the target population is itself a study based on a population sample, so the population quantities that we treat as true are in fact uncertain. Unusually, the target population is also much smaller that the UK Biobank imaging cohort, adding additional uncertainty to the analysis. Furthermore, bias that we attribute here to preferential selection may be from another source, like measurement error. For example, the HSE and UK Biobank have different questionnaires, and, for example as respondents to report education level in slightly different ways. While care has been taken here to standardize the responses across sources, there is likely some lingering discrepancy.

Second, the crucial assumption underlying all adjustment procedures tested here is that we have correctly identified an admissible set. While we introduce the concept of an admissible set and criteria necessary for recovering from selection bias, we do not actually identify such a set in the UK Biobank. We also limit our analysis only to auxiliary variables for which we have external population data readily available, when that is not a requirement for recovery in all cases. Furthermore, there is a wealth of data available in the UK Biobank that we fail to leverage, perhaps most obviously spatial data.

The third major limitation involves our treatment, or lack thereof, of variance of estimators or statistical significance of results. We only evaluate relative performance based on visual assessments, not based on statistical tests, so cannot make any claims about a method having significantly better performance than another.

Lastly, we only perform simulation studies in which the missingness mechanism is static. Adjustment methods that performed well could be particularly suited to the characteristics of the missingness used here.

There are many possible avenues for future research. First, we could continue to explore and refine various adjustment procedures. For example, we could adapt calibration and raking to better handle larger numbers of variables. We could also expand our analysis to consider methods that directly model the outcome of interest, like Multilevel Regression and Poststratification, or MRP park2004bayesian. We could explore tuning parameters for the BART and LASSOs.

Second, we could incorporate additional auxiliary variables, like spatial data, into weighting procedures. As discussed, many weighting procedures suffer from an inability to handle a large number of auxiliary variables, requiring the researcher to manually select variables and interactions they think will be important

Another direction for future research is incorporating computation time into overall analysis of estimators. BART, for example, performs well, but also takes much more computation time than post-stratification, for example.

## Data Availability

Data can be accessed through the UK Biobank.

https://www.ukbiobank.ac.uk/

## 6 Code and Data Availability

All code is publicly available at https://github.com/vcbradley/ukb-selection-bias.

This analysis sources data from the UK Biobank and the 2016 Health Survey for England (HSE). UK Biobank data is publicly accessible upon approval of an application through https://bbams.ndph.ox.ac.uk/ams/signup. HSE data was accessed via the UK Data Service at https://ukdataservice.ac.uk.

The UK Biobank is the selection-biased data that we would like to adjust, and the 2016 HSE is our target population. As we aim to directly compare demographic and health metrics from the UK Biobank to those from the HSE, we recode relevant variables to match across sources as closely as possible. For example, this may include collapsing levels of a certain variable in the UK Biobank because the corresponding variable was collected at a higher level of aggregation in the HSE. We will describe a few such cases in the rest of the section, but full recodes can be found in the paper’s GitHub repository.

## 7 Ackowledgements

Computation used the BMRC facility, a joint development between the Wellcome Centre for Human Genetics and the Big Data Institute supported by the NIHR Oxford BRC. The views expressed are those of the author(s) and not necessarily those of the NHS, the NIHR or the Department of Health.

### Appendix

#### A Data

Demographic variables, like household income and highest education level, were collected once at baseline, and again at the time of the imaging assessment. As we are interested in correcting potential selection bias in brain MRI data with auxiliary demographic variables, we take the value collected at the time of imaging wherever available. If missing, we take the next most recent observation.

Some variables, like income and home ownership, contain observations where the respondent refused to respond or was unsure of the correct response. We code those cases as “Do not know/Refused” and do not impute response values, and do not exclude them from the data, as doing so may introduce additional bias. The HSE also contains “Do not know/Refused” values for these variables, so there is a subset for comparison, though the pattern of refusals may be different across the two studies.

Some variables, like age and ethnicity, are coded in multiple ways - once at the most granular level for use in simulation studies, and once at a higher level of aggregation to match the way the information is collected in the HSE. Age, for example, is collected in the UK Biobank as a combination of month and birth year. We impute continuous age as the “age in years” from the 15^th^ of the observed birth month and the date of the imaging appointment. In the simulation studies, we use a continuous version of age, however, the HSE only reports age in 5-year increments. Therefore, we create a discrete version of UK Biobank age to match the HSE variable which is used to weight the Biobank data. In the UK Biobank, subethnicity, like “White Irish” and “Black African” is collected in addition to the larger ethnicity categories: white, black, Asian, mixed, other, and no response. However only the major categories were collected by the HSE. Therefore, we code ethnicity two ways: one at the most granular level to use in simulation studies and one at the higher level of aggregation for weighting to the HSE.

We also consider a small selection of health outcomes:

- **Smoking status** (4 categories): current, previous, never, do not know/refused
- **BMI category** (5 categories): underweight (*<* 18.5), healthy (18.5 − 24.9), overweight (25 − 29.9), obese (*>* 29.9), do not know/refused
- **Ever diagnosed with diabetes by a doctor** (3 categories): yes, no, do not know/refused
- **Ever diagnosed with high blood pressure** (3 categories): yes, no, do not know/refused

These variables were selected because the HSE collects similar outcomes, giving high-quality population prevalence estimates for comparison. These health outcomes were not used in the simulation studies, or to weight the UK Biobank data, but serve as benchmarks for how much healthy volunteer selection bias exists in the UK Biobank and how well weighting methods are able to adjust for it.

##### A.0.1 UK Biobank Imaging Data

In the UK Biobank, there are 20,827 subjects that have a recorded MRI brain volume, and will serve as the population for these simulations. We observe the following demographic data for each subject:

- **Age** measured at time of imaging appointment
  **–** continuous: 40 - 79
  **–** squared: 40^2^ - 79^2^
  **–** discrete (7 categories): 45-49, 50-54, 55-59, 60-64, 65-69, 70-74, 75-79
- **Sex** (2 categories): Female, Male
- **Ethnicity** (12 categories): White, White Irish, White other, Asian Indian, Asian Pakistani, Asian Bangladeshi, Asian Other, Black Caribbean, Black African, Mixed, Other, do not know/refused
- **Employment** (8 categories): employed, retired, homemaker, disabled, volunteer, student, unemployed, do not know/refused
- **Occupation** (SOC2010, 11 categories): manager, professional, associate professional, administrative, skilled trades, personal service, sales or customer service, industrial, elementary, unemployed, do not know/refused
- **Highest level of education** (6 categories): college or above (including professional degree), A-levels (or equivalent), O-levels or CSEs, vocational or other, none, do not know/refused
- **Household income** (£1000s, 6 categories): Under £18, £18-£31, £31-£52, £52-£100, Over £100, do not know/refused
- **Household size** (6 categories): 1, 2, 3, 4, 5 or more, do not know/refused
- **Home ownership** (6 categories): Own outright, own with a mortgage, rent from LA, rent from a private landlord, rent free, do not know/refused
- **Home type** (3 categories): house, flat/apartment/temporary accommodation, do not know/refused

#### B Adjustment methods

##### B.1 Post-stratification

One of the main disadvantages of post-stratification is the need for the researcher to select variables with which to strata for weighting that capture as much of the missingness mechanism as possible without resulting in tiny cells that contain few or no observations. This is typically a manual process that relies on domain knowledge, however, that is impractical in this setting. We use a random forest to simulate the domain knowledge typically used for variable selection. Specifically, we use a random forest (from the `randomForest` package in R) to predict the vector of selection indicators **s** using the discrete *Z*_*k*_ in their categorical (not binary indicator) forms. We exclude the continuous *Z*_*k*_ as post-stratification can only use discrete variables, and implicitly considers variable interactions, so there is no need to explicitly specify them in the random forest.

The random forest generates a measure of variable importance. Beginning with the variable deemed most important, we add on additional variables of decreasing importance until the joint distribution of the selected variables creates cells that have no observations in the sample and make up, in aggregate, no more than 1% of the population. For example, consider if the two most important variables were age buckets and gender and no cells based on the interaction of the two variables were empty in the sample. We would add the next important variable, say income, and re-calculate the number of observations in each cell of the joint distribution of all 3 variables. Say that there are 2 cells for which we lack sample observations, but combined, they make up only 0.5% of the population. This is acceptable, and we move on to consider the next most-important variable, employment status. The joint distribution of all 4 variables contains 10 empty sample cells, which make up 1.5% of the population, which is over out threshold. Therefore, we eliminate the 4^th^ variable and proceed to post-stratification with the first three.

#### B.2 Calibration

Calibration minimizes the distance between prior weights, set to 1 here, and new weights that satisfy a set of constraints. Constraints are defined as functions of auxiliary variables, often population totals. Here we consider 3 sets of constraints:

- Population size in the form of 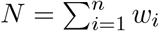
- Sums of continuous variables (age and age^2^) in the form of 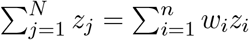
- Counts across levels of the 10 categorical variables. For this constraint, we transform categorical variable *Z*_*k*_ that has *L*_*k*_ levels into *L*_*k*_ − 1 binary indicator variables, dropping one level per variable to avoid collinearity. Constraints take the form

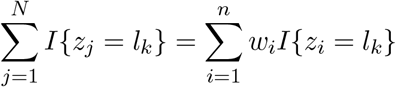

for all *l* ∈ (1, …, *L*_*k*_ −1) levels of *Z*_*k*_, and all *Z*_*k*_ in the *k* ∈ (1, …, 10) categorical variables.

In total, there are (1 × population total + 2 × age totals + 6 × age categories + 1 × sex + 11 × ethnicity, 7×employment+10×occupation+5×education+5×income+5×household size+5× home ownership + 2 × home type) = 60 constraints to consider in each calibration estimation. With so many constraints, calibration can perform poorly - either by producing extreme weights, or by failing to converge altogether. In order to prevent this, we eliminate constraints that apply to levels of discrete variables that make up less than 2% of the sample or the population. By eliminating these constraints, we effectively pool these small levels with the reference level for that categorical variable, which has been chosen somewhat arbitrarily. Generally in a real application setting, the researcher would manually pool small strata based on domain knowledge.

The second step we take to aid convergence is calibrating in two stages. We split the constraints into two groups based on the number of observations in the population that are represented by that level, and calibrate the sample first with the constraints for smaller population subgroups, then use the resulting weight as the prior weight for a second round of calibration with the larger constraints. We calibrate the smaller groups first so that the final weighted sample exhibits less overall error in marginal distributions of auxiliary variables. This approach is ad-hoc, and should be tested against other methods for dealing with a large number of constraints in future research.

The last step we take to aid convergence in calibration is the specification of algorithm parameters - namely the tolerance threshold and the maximum number of iterations that the algorithm will be allowed. The tolerance threshold *ϵ* is the threshold that determines when the weighted sample total matches the population total:

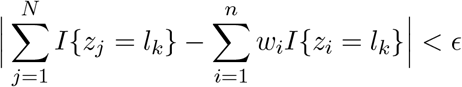

By default, *ϵ* is set to 1*e* − 7 * *N*, however we raise this threshold to *ϵ* = 0.0003 * *N*. This is primarily based on anecdotal observations of cases when calibration failed to converge despite other precautions taken. Further research should explore performance of calibration weights as a function of this tolerance. Lastly, the maximum number of iterations was increased from the default of 50 to 5000.

We used the `calibrate` function from the `survey` package in R to fit calibration weights.

##### B.3 Raking

As discussed in Section **??**, raking is a specific form of calibration in which the constraints are marginal distributions of categorical variables. As in calibration, raking will fail to converge when there are a large number of constraints or when constraints include discrete variables with levels representing small population subgroups. In order to avoid this, we eliminated members of the population that were absent in the sample, as is standard practice in poststratification. For example, if the sample contained no observations with ages between 40 and 45, we eliminated all members of the population between those ages, and calculated target population marginal distributions based on the remaining members of the population. This clearly presents a problem if we are forced to drop small, but potentially important, population subgroups to fit rake weights. In most real applications, the researcher would pool levels of the population that were missing from the sample with other levels based on domain knowledge. However this is a manual process, and a large drawback of raking, which we would like this simulation to capture.

We used the `rake` algorithm in the `survey` package in R to fit rake weights. This implementation allows for the specification of a tolerance threshold *ϵ* and of a maximum number of iterations. As in calibration, we set the tolerance threshold to be 0.0003, and the maximum number of iterations to be 5000.

##### B.4 Raking with LASSO variable selection

This method approaches adjustment as a variable selection problem. Raking can consider a large set of auxiliary variables, while post-stratification quickly becomes impossible when more than a few auxiliary variables are considered. On the other hand, post-stratification can account for interactions in the missingness mechanism, while raking only adjusts the marginal distributions of the auxiliary variables. This method attempts to leverage the advantages of each raking and post-stratification by adjusting on the minimum set of interactions significantly related to the outcome, in this case total brain volume, or probability of selection. LASSOs are used to select significant predictors of brain volume or selection, and then the sample is raked only to the marginal distributions of those variables. We excluded age and age^2^ from consideration, as we cannot rake to the population margins of continuous variables. This left 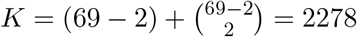 possible predictors.

For the LASSO predicting selection, we use all 20,827 observations in the population and predict the binary outcome *s*_*j*_ which is 1 if the *j*^th^ observation was selected in the sample, and 0 otherwise. For the outcome LASSO, we restrict the training data to the sample and assume that the outcome, brain volume, is normally distributed conditional on covariates **X**. We use `cv.glmnet` from the `glmnet` package in R, and 5-fold cross-validation.

In practice, there are a few challenges to address. First, any variable selected by one of the LASSOs will be used in raking, so there must be enough observations of each level in both the sample and population. We set that threshold to be 1%, thus eliminating any variables that create population or sample cells below this threshold.

Second, in order to decrease the computation required to fit each LASSO, we reduce the number of *λ* penalty values considered. By default, `cv.glmnet` considers 100 values of *λ* from *λ*_min_ to *λ*_max_, equally-spaced on the log scale. *λ*_max_ is set such that when *λ > λ*_max_, all coefficients are 0. Friedman et al. (2010) show that *β*_*k*_ will be 0 if 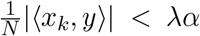, and therefore all coefficients will be 0 when *Nλ*_max_ = max_*k*_ |⟨*x*_*k*_, *y*⟩|. *λ*_min_ is set to 0.001*λ*_max_. We take a similar approach but consider only 20 values between *λ*_min_ and *λ*_max_ instead of 100. The final set of variables was selected using the largest *λ* such that the cross-validated error lies within 1 standard deviation of the minimum cross-validated error. This criterion is often used to prevent overfitting (Friedman et al., 2010).

Last, the LASSOs frequently identified 30 or more significant variables, which, when considered all at once, caused the raking algorithm to fail to converge. To prevent this, we capped the number of raking variables at 50 and then raked using sets of 20 variables at a time, from least important to most important. On each raking iteration, the weights from the previous iteration were used as the prior weights. Variable importance was determined first by whether the variable appeared in one or both LASSOs, and second by the relative importance of the variable within the LASSO. Due to the difference in the scale of the outcomes across the two LASSOs, we relied on relative coefficient size rather than absolute coefficient size.

Once raking variables and groups were identified, we implemented raking using the same settings as the straight raking approach described above.

##### B.5 Logistic regression

Logistic regression for selection bias adjustment takes a different approach than the previous methods by attempting to directly estimate the probability of selection instead of attempting to make the sample margins match the population margins. In fact, this method makes no attempt to match population margins.

We use a logistic regression LASSO for variable selection. Similarly to the LASSOs used in the previous method, we used `cv.glmnet` with 5-fold cross-validation and custom values of *λ*. The optimal *λ* was selected using the 1 standard deviation criterion, and the model was then re-fit using only significant predictors and no penalty parameter to avoid coefficient shrinkage in the final predictions. Occasionally, the LASSO would fail to select any significant variables. In that case, a simple logistic regression was fit using all 69 first-order predictors and no interactions.

The weights from the resulting logistic regression are 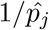 where 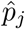 is the modeled probability of selection.

##### B.6 BART and raking

BART and raking is similar to the previous method in that it attempts to directly estimate the probability of selection. The additional raking step is then ensures that sample marginal distributions of key variables match those of the population.

We use the `BayesTree` package in R to fit the BART on all 69 first-order terms, including age and age^2^. BARTs naturally consider non-linearities and interactions between predictors, so there is no need to manually pre-specify them. We fit a categorical BART with 25 trees, and the estimate the probability of selection for each subject with the mean of 1000 samples from the posterior of the model.

The BART-estimated probabilities serve as our prior probabilities in raking. To avoid over-raking, and potentially increasing the variance unnecessarily, we select a subset of variables for raking. The `BayesTree` implementation of BART does not have a variable importance metric, so we fit a simple random forest from the `randomForest` package to the same data using the 8 categorical covariates instead of the 67 binary covariates, and select the top 5 most important variables from that model for raking. Raking settings were the same as those used for simple raking, described above.

###### Other considerations

For all methods, weights were re-scaled to have mean 1 to simplify comparison across methods. Occasionally methods would fail to converge despite precautions taken. In that case, a weight of 1 was assigned to all sampled units.

##### B.7 Application to the UK Biobank

With an understanding of the relative benefits of various adjustment procedures from the previous section, we will then apply them to the actual UK Biobank imaging data. We will attempt to estimate the following population characteristics:

- **Prevalence** of smoking
- **Prevalence** of obesity
- **Prevalence** of ApoE e4/e4 phenotype
- **Population mean** total brain volume
- **Association** between age and total brain volume

This list of outcomes represent two broad categories of outcomes with different levels of strictness of conditions for recovery that are both of interest in studies like the UK Biobank: prevalence P(**Y**) and association P(**Y**|**X**). Second, these outcomes have been widely studied, so the literature provides a strong benchmark by which to evaluate our adjustment procedures (Brandon et al., 2018; Fotenos et al., 2008).

To estimate these outcomes, we will apply each of the weighting methods under consideration to the UK Biobank imaging cohort using the 2016 Health Survey for England (HSE) as our target population to define population totals of auxiliary variables. Then, we will calculate weighted estimators of each quantity. As in the simulation study, the association between total brain volume and age will be calculated using weighted linear regressions.

We will calculate the design effect of each set of weights, and compare estimates to population quantities from the 2016 HSE.

#### C Simulation

##### C.1 Probability of missingness

The simulation requires generation of a missingness mechanism, or a linear combination of demographic variables **Z** and randomly-drawn coefficients ***β*** that is used to estimate a probability of selection *p* for each subject in the population, *p*_*j*_ = logit(***β*Z**_*j*_).

We generate a single set of coefficients ***β***, which we would like to be 1) perfectly recoverable, 2) create significant bias in estimates of outcomes and 3) be realistically complex.

In order to ensure the first criterion, probability of missingness is a function only of variables **Z** that can be considered by the weighting procedures (listed in the previous section). For the second criterion, we always include age in the missingness function, which is well-known to be correlated with brain volume. In the UK Biobank imaging data, age and brain volume have a correlation of -0.55.

The third criterion stems from the result shown in Equation **??**. We know that if we observe all variables **Z** that d-separate **Y** and *S*, and can estimate ŷ in each cell defined by the joint distribution of **Z**, we will always be able to unbiasedly recover the quantity of interest using post-stratification, and post-stratification will dominate other methods (Caughey and Hartman, 2017). However in most practical applications, **Z** is large or includes a continuous variable, making it impossible to estimate ŷ in all cells formed by the joint distribution of **Z**.

We seek to evaluate adjustment methods under realistic conditions, so would like **Z** to be large and contain continuous variables. Age will always be included in order to induce bias (criterion 2). We also ensure that **Z** is complex by considering each level of each discrete variables separately by coding all categorical variables as a set of indicator variables, without removing a default reference value. For example, home type is coded as 3 variables, each corresponding to one of the levels (example shown in Table 5).

**Table 5:** Example of variable coding for simulating the probability of missingness.

| **Home type** | **house** | **flat_or_apartment** | **refused** |
| --- | --- | --- | --- |
| House | 1 | 0 | 0 |
| Flat or apartment | 0 | 1 | 0 |
| Do not know/refused | 0 | 0 | 1 |

This clearly creates collinearity between predictors, and we will rely on the sparsity of the simulated ***β*** to ensure that not all levels of a single categorical variables are included in a single function. Furthermore, even if by chance all levels of a variable were included, since we are simply simulating a function for *p* and not estimating it in a regression, one of the levels could be considered an intercept term.

Real missingness mechanisms are often non-linear. We introduce non-linearity here by including *age*^2^ in **Z** and by considering all two-way interactions of demographic variables. We have (age, age^2^, 7 × age categories, 2 × sex, 12 × ethnicity, 8 × employment, 11 × occupation, 6 × education, 6 × income, 6 × household size, 6 × home ownership, 3 × home type) = 69 first-order predictors. Considering two-way interactions introduces an additional 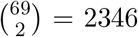 predictors, for a total of *K* = 2415 possible predictors. While this may seem extensive, we have only considered a small fraction of the variables available in the UK Biobank, and only one type of non-linearity.

We use a spike-and-slab distribution to simulate the missingness coefficient *β*_*k*_ for all *k* ∈ (1, …, *K*) predictors. The spike and slab distribution is a mixture model, using a Bernoulli distribution to model the probability that random variable is non-zero (the “spike”) and a Normal distribution to model the value of the variable given that it is different from 0 (the “slab”). The parameters of the distribution are *λ*_*k*_, the probability that *β*_*k*_ is non-zero, and *µ*_*k*_ and 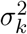 the mean and variance of the Normal distribution that *β*_*k*_ follows if non-zero. The hyperparameter *α*_*k*_ indicates if *β*_*k*_ is non-zero.

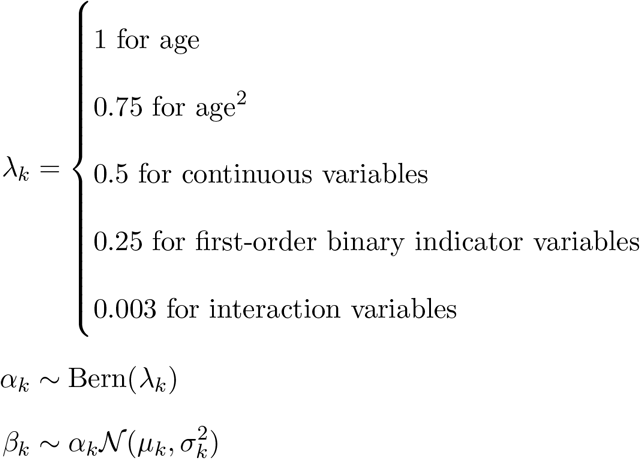

The sampled values of ***β*** used in the simulation study are given in Table 6 in the Appendix.

**Table 6:** Missingness coefficients used in the simulation study.

| $Z$ | $\beta_z$ |
| --- | --- |
| age | 2.2362584 |
| demo_sexMale | -2.3681210 |
| demo_age_bucket60 to 64 | -0.5221229 |
| demo_ethnicity_full99-DNK/Refused | -0.3310816 |
| demo_empl_retired | -0.6548054 |
| demo_empl_unemployed | 1.0396497 |
| demo_empl_student | -1.2736848 |
| demo_occupation04-admin | 1.0748084 |
| demo_occupation99-DNK/Refused | -0.6608140 |
| demo_educ_collegeplus | -0.7038866 |
| demo_educ_highest_full02-A Levels | 1.8571014 |
| demo_educ_highest_full03-O Levels | -0.6431069 |
| demo_educ_highest_full07-None | 0.5751345 |
| demo_income_bucket04-52k to 100k | 2.3920356 |
| demo_hh_size4 | 0.5362810 |
| demo_hh_ownrent02-Own with mortgage | -0.6211912 |
| demo_hh_ownrent99-DNK/Refused | 0.8182660 |
| age_sq | 0.0915620 |
| age:demo_ethnicity_full04-Mixed | -1.6169544 |
| demo_age_bucket50 to 54:demo_empl_disabled | -0.7442275 |
| demo_ethnicity_full05-Asian Indian:demo_empl_volunteer | 2.0003787 |
| demo_ethnicity_full06-Asian Bangladeshi:demo_hh_ownrent06-Rent free | -2.2941626 |
| demo_empl_retired:demo_hh_ownrent06-Rent free | -1.1010164 |
| demo_empl_disabled:demo_occupation07-sales customer service | 0.7285372 |
| demo_empl_disabled:demo_hh_ownrent03-Rent from LA | 1.5001765 |
| demo_occupation08-industrial:demo_hh_ownrent04-Rent private | 1.7046601 |

As described in Algorithm 1, once the ***β*** = (*β*_1_, …, *β*_*K*_) have been sampled, we calculate *p*_*j*_ for all *j* ∈ (1, …, *N*) as *p*_*j*_ = logit(**Z**_*j*_***β***). Then, holding the sample size *n*_*sim*_ = *Nπ*_*obs*_ fixed, we draw a sample from the population. Let *s*_*j*_ be the indicator for the *j*^th^ person being sampled, then *s*_*j*_ ∼ Bern(*p*_*j*_|*n*_*sim*_). We consider a range of proportions observed *π*_*obs*_ from 0.01 to 0.25 in order to evaluate how the performance of adjustment procedures varies with the amount of data available.

##### C.2 Distribution Bias

**Distribution bias** measures how closely they estimate the population marginal distributions of auxiliary variables used in weighting. For example, consider an auxiliary variable *Z* with levels *l* = (1, …, *L*_*z*_), and corresponding population totals 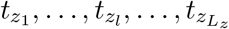, where 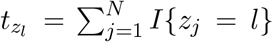. Then, the proportion of the population made up by level *l* of *Z* is 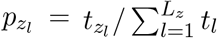. The weighted estimator for 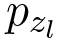 is

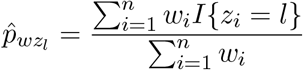

for a sample of units *i* ∈ (1, …, *n*).

We will define the distribution bias (DB) for a set of weights to be the sum of the squared bias of 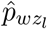 across all levels of all auxiliary variables:

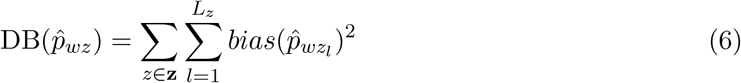

#### D Additional Figures and Tables

**Figure 10:**
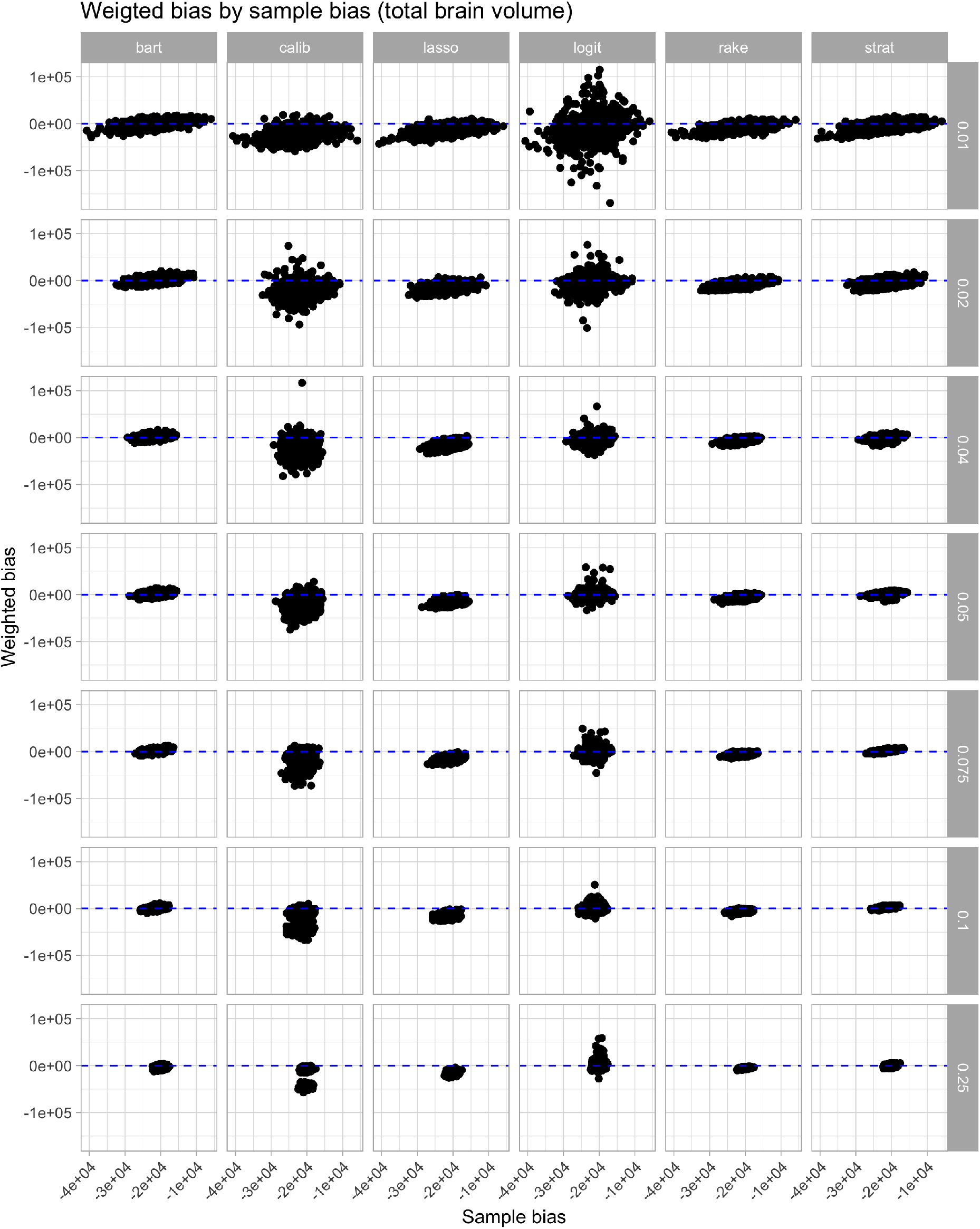
Bias in total brain volume. The x-axis shows the actual selection bias in the sample, the y-axis shows the remaining bias once each adjustment procedure was applied.

**Figure 11:**
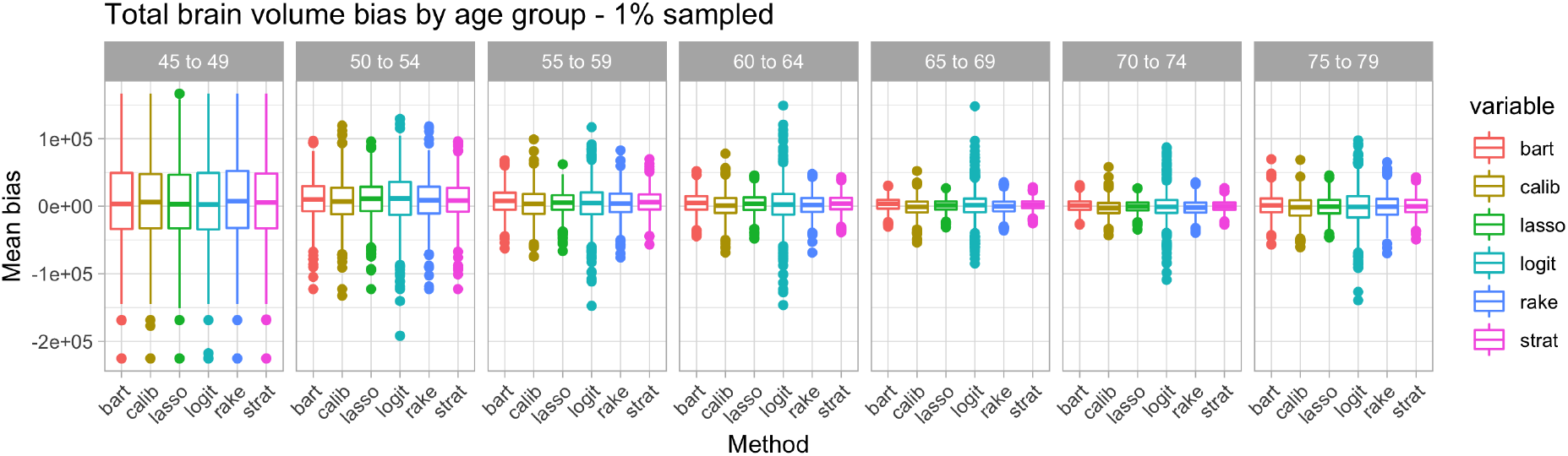
Bias of total brain volume by proportion sampled

**Figure 12:**
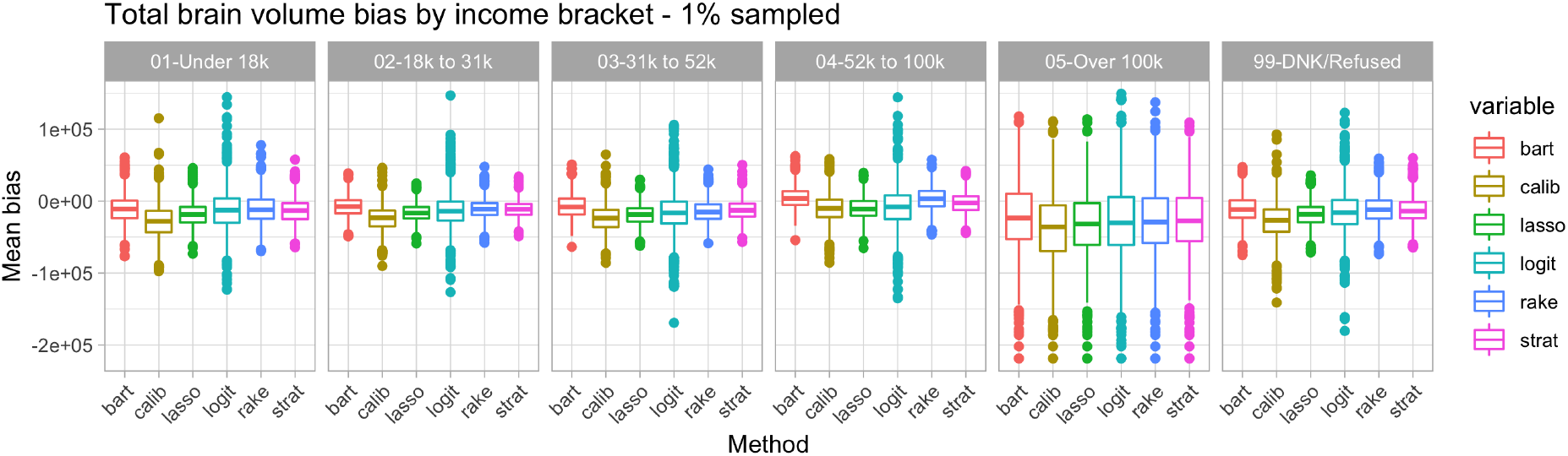
Log median design effect of weights by average bias in total brain volume

